# Genetic and Epigenetic Foundations of Childhood Internalizing and Externalizing Problems and their Co-occurrence

**DOI:** 10.1101/2025.05.24.25328240

**Authors:** M. Bekkhus, S. Tsotsi, CM. Page, NO. Czajkowski, W. Liu, E. Røysamb, Nes R. Bang, A. Oftedal, R. Lyle, L. Hannigan, OA. Andreassen, Y. Wang

## Abstract

Childhood behavioural problems pose long-term mental health risks, yet the biological basis of co-occurring internalizing and externalizing symptoms remains unclear. In a cohort of 40,212 children, we integrated genome-wide association studies (GWAS), polygenic scores (PGS), and DNA methylation profiling (EWAS) to examine the genetic and epigenetic underpinnings of internalizing, externalizing, and co-occurring problems. All outcomes showed modest SNP-based heritability (h² = 0.02–0.19, FDR < 0.05). GWAS identified a significant locus for co-occurring problems at age five (rs73161798, *p* = 3.26 × 10⁻⁸), near *USP12*, a gene implicated in neuroplasticity and risk-taking. PGS for co-occurring symptoms was associated with higher neuroticism and lower cognitive ability. DNA methylation explained substantial variance in co-occurring problems at ages three (R² = 0.19) and five (R² = 0.62), but not in single-domain symptoms. EWAS revealed 101 significant sites (p < 10⁻⁷) linked to neurodevelopmental, immune, and metabolic pathways. These findings suggest that co-occurring behavioural problems have a distinct, age-sensitive biological architecture.

## INTRODUCTION

Child behavioral problems, encompassing both externalizing and internalizing difficulties (such as impulsivity, hyperactivity, and aggression) and internalizing difficulties (such as anxiety, depression, and emotional distress)^1^—are highly prevalent and have a substantial impact on children’s cognitive and social development. These problems also pose long-term risks for both mental and physical health^1,2^. Although internalizing and externalizing problems have traditionally been considered distinct domains, growing evidence suggests they frequently co-occur, indicating potentially shared underlying mechanisms^2–4^.

The co-occurrence of internalizing and externalizing problems complicates both diagnosis and intervention, as children with comorbid problems often follow different developmental trajectories and are at higher risk for severe and persistent psychiatric disorders compared to those with isolated symptoms^5–8^. Indeed, accordingly research has introduced the “p-factor” hypothesis, which suggests that a general liability to psychopathology underlies various mental health problems, rather than each diagnosis being entirely distinct^5,9,10^. Specifically, this model proposes that genetic and environmental influences on early-life behavior contribute to broad psychiatric vulnerability, rather than being unique to specific disorders.

A person-centered approach, such as the one used by Tsotsi et al. (2023), has identified distinct co-occurring profiles of internalizing and externalizing behaviors, particularly highlighting the overlap between internalizing difficulties and aggression. This approach assumes that individuals vary in the level of internalizing and externalizing symptoms, allowing for more accurate representation of individuals that traditional methods such as cut-off scores on a given measure. Children exhibiting these co-occurrent symptoms have been found to be at a greater risk for persistent and severe mental health issues later in life, including depression, anxiety, and conduct-related disorders^11,12^. However, while co-occurring behavioral problems may share common risk factors, they may also have unique underlying causes and outcomes, emphasizing the importance of distinguishing shared genetic influences from disorder-specific genetic and environmental factors. Despite growing evidence for genetic contributions to child behavior^13^, few studies have explored how genetic and epigenetic mechanisms contribute to the development of co-occurring behavioral problems in early childhood. Addressing this gap is crucial for improving early detection, intervention, and prevention strategies.

Psychiatric genetics has made major advances in recent years, identifying common genetic variants associated with psychiatric disorders^14–18^, as well as common and distinct genetic underpinnings of externalizing and internalizing behavior^19^. While GWAS findings suggest both shared and distinct genetic influences across these domains, the genetic architecture of co-occurring childhood behavioral problems remains largely unknown^20^. Emerging evidence indicates that co-occurrence is partly driven by shared genetic risk, particularly for traits like ADHD and lower cognitive ability^21^, though differentiation between subtypes remains limited. Similarly, studies of co-occurring depression and conduct disorder suggest a strong influence of shared genetic liability, consistent with transdiagnostic models of psychopathology^22^. These findings underscore the importance of studying co-occurrence as a genetically meaningful construct, not merely the sum of internalizing and externalizing symptoms.

In addition to genetics, epigenetic modifications such as DNA methylation (DNAm) may play a role in behavioral regulation. DNAm reflects both genetic predisposition and environmental influences, making it a critical factor in understanding behavioral development^23^. Previous epigenome-wide association studies Epigenome-wide association studies (EWAS) have linked DNAm to psychiatric traits, including schizophrenia^24^, attention deficit hyperactivity disorder (ADHD)^25^, tobacco smoking^26^, and externalizing and internalizing problems^27^. However, few studies have integrated both genetic and epigenetic data to investigate the origins of co-occurring childhood behavioral problems.

This study leverages a large Norwegian birth cohort (MoBa)^28^ to investigate genetic and epigenetic influences on early behavioral problems. Using genome-wide association studies (GWAS), polygenic risk scores (PGS), EWAS, weighted correlation network analysis, and Mendelian randomization (MR), we examine whether co-occurring behavioral problems have a distinct genetic and epigenetic basis compared to internalizing or externalizing problems alone. Thus, in this study we test the model as illustrated in **Fig.1A** and aim to (1) explore the genetic and epigenetic variations in behavior outcomes by identifying unique genetic variants and differentially methylated positions (DMPs) associated with externalizing and internalizing and co-occurring behavioral outcomes, (2) assess whether DNAm variations contribute causally to these behavioral outcomes, and (3) explore shared genetic architecture with neuropsychiatric and cognitive traits.

**Figure 1.**
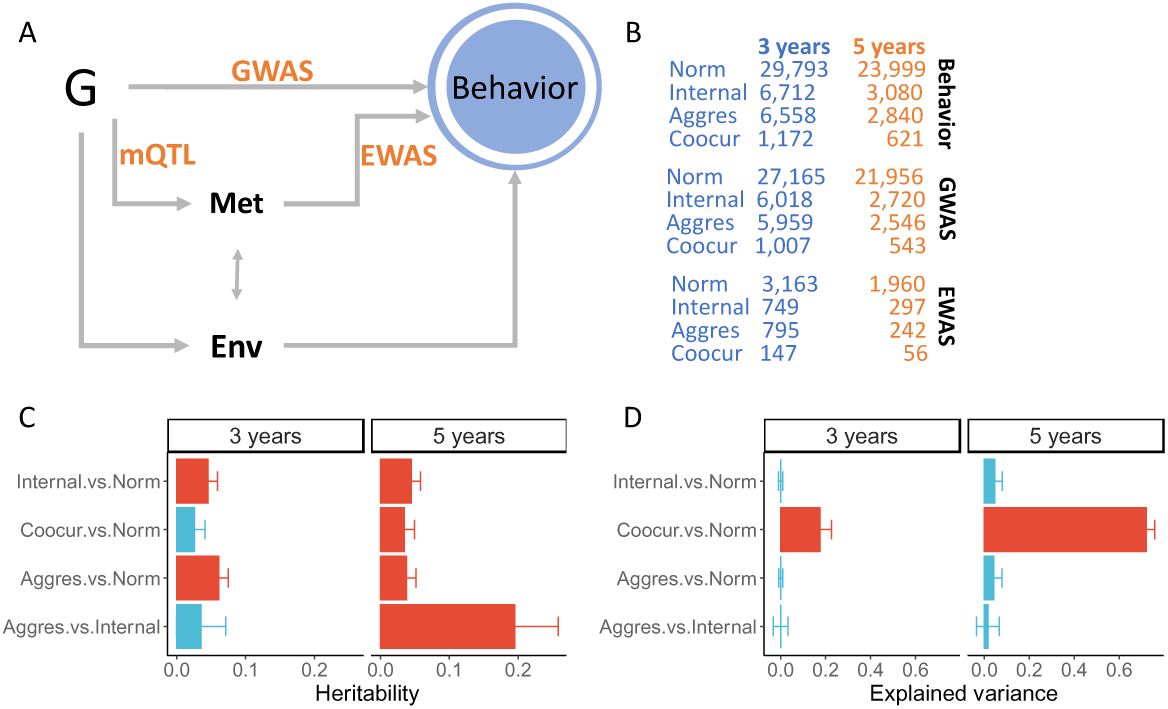
The study scheme and explained phenotype variance by genomic-wide SNPs and epigenome-wide methylation levels. A. Major analysis steps; and B. Sample sizes for different analysis steps. Genome-wide association studies were performed from common genetic variants (G) to child behavioral problems (Behavior). Epigenome-wide association studies were performed from DNA methylation levels in cord blood to child behavioral problems (Behavior). Methylation quantitative trait loci (mQTL) analyses were performed from common genetic variants to significant methylation sites. In utero environmental factors (Env) were accounted for in the DNA methylation analyses. C. SNP-heritability for the contrasts between child behavioral problems and normal behavior is shown on the y-axis. D. The proportions of variation in the contrasts, as in (C), that were explained by DNA methylation variation are shown on the y-axis. Datasets from the two ages are shown separately in the headers. Standard errors of the estimates are also presented. Norm: normal behavior; Internal: primarily internalizing problems; Aggres: aggressive behavior; Coocur: co-occurring internalizing and aggressive behavior.

By integrating genetics and epigenetics, our study provides new insights into the biological underpinnings of childhood behavioral problems and their potential long-term mental health risks. Understanding these mechanisms may inform early intervention strategies and improve risk prediction for psychiatric disorders.

## RESULTS

### Child Behavior Problems Explained by Genome and Epigenome Variations

To estimate the genetic contribution to childhood behavioral outcomes, we calculated SNP-based heritability (SNP-h²) using GCTA (**Methods**). At age three, we observed significant SNP-h² estimates for internalizing problems (SNP-h² = 0.05, SE = 0.01, p = 3.5 × 10⁻⁴) and aggression, a subgroup of externalizing behavior (SNP-h² = 0.06, SE = 0.01, p = 2.02 × 10⁻⁶) (**Fig. 1C)**. The co-occurring behavioral problem showed a nominally significant heritability estimate (SNP-h²= 0.03, p = 0.048) but did not survive multiple testing correction. By age five, all four behavioral phenotypes exhibited significant SNP-h² estimates, with aggression showing the highest heritability (SNP-h² = 0.19, SE = 0.06, p = 9.45 × 10⁻⁴), but only when it is compared to internalizing problems (**Fig. 1C**; see **Methods**). Full heritability estimates are presented in **Supplementary Tables S1** and **S2**.

In parallel, to estimate the epigenetic contributions to behavioral problems, we estimated the proportion of behavioral variance explained by epigenome-wide DNAm in umbilical cord blood. Interestingly, only the co-occurring problem showed significant DNAm-associated variance at both ages (three years: variance explained = 0.17, SE = 0.05; five years: variance explained = 0.82, SE = 0.03) (**Fig. 1D**). These results suggest that co-occurring behavioral problems may have a strong epigenetic component, whereas internalizing and externalizing problems appear more associated with genotypes. Full estimates for all traits are provided in **Supplementary Tables S3** and **S4**.

### Genetic Variants Associated with Child Behavioral Problems

To identify specific genetic variants, we performed GWAS for each behavioral phenotype at ages three and five (**Fig. 1A**). Among the eight GWAS performed (four per age), only the co-occurring problem at age five yielded a genome-wide significant locus (**Figs. S1 and S3**). This signal (p = 3.26 × 10⁻⁸) was located on chromosome 13, led by SNP rs73161798, which resides in an intergenic region near ubiquitin-specific peptidase 12 (*USP12*) (**Fig. 2A**). USP12 has been previously linked to risk-taking behavior and neuroplasticity ^29,30^. Quantile-quantile (QQ) plots (**Suppl. Figs. S2** and **S4**) and lambda inflation factors (range: 1.002–1.012) indicated no substantial genomic inflation, supporting the robustness of this finding.

**Figure 2.**
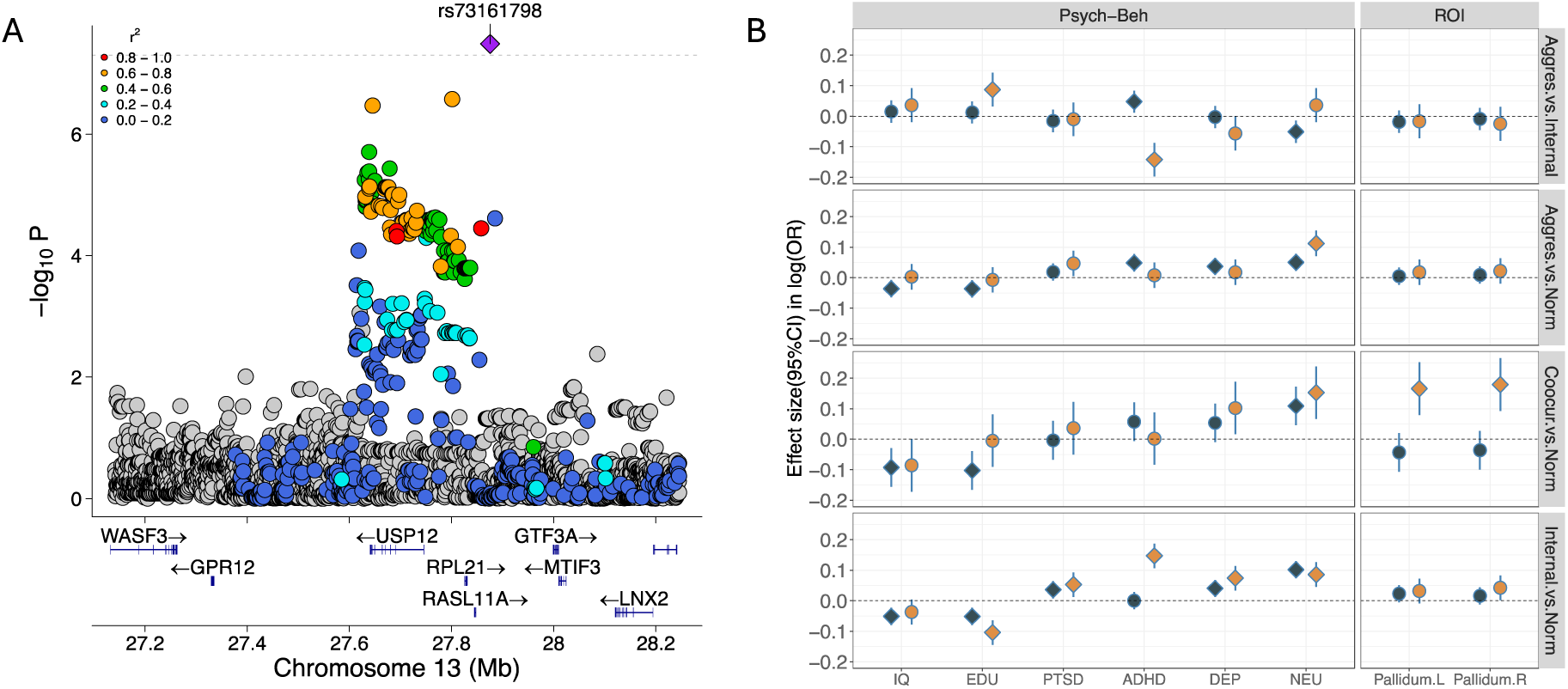
Genetic influence on child behavior problem. A. The locus associated with the co-occurring problem in contrast to normal behavior. The GWAS lead SNP is highlighted; the association strength (−log10 p-value) is shown on the y-axis, and the genomic location is shown on the x-axis. Nearby genes are displayed in the bottom panel. B. Effects, presented on a log odds ratio scale, of polygenic scores for eight traits (x-axis) on child behavior problems. Results for the two ages are distinguished by color (blue: three years; red: five years). The 95% confidence intervals are shown, and statistically significant associations (FDR ≤ 0.05) are indicated by diamond shapes. Traits and disorders are grouped into psychiatric-behavioral (Psychi-Beh) and brain imaging regions of interest (ROI). Abbreviations: IQ – intelligence quotient;EDU – educational attainment;PTSD – post-traumatic stress disorder;ADHD – attention deficit/hyperactivity disorder;DEP – depressive symptoms; NEU – neuroticism; Pallidum.L/R – volumes of the brain pallidum on the left and right sides, respectively; Norm – normal behavior; Internal – primarily internalizing problems; Aggres – aggressive behavior; Coocur – co-occurring internalizing and aggressive behavior.

Given that SNP-h² estimates and GWAS results indicated a polygenic nature for child behavioral problems, we PGS analyses to investigate genetic relationships between childhood behaviors and common psychiatric disorders, cognitive traits, and brain structure (**Methods**). After multiple testing correction, we identified six psychiatric/cognitive traits significantly associated with at least one childhood behavioral problem (**Fig. 2B**, **Suppl. Tables S5** and **S6**). At age three, higher risk for internalizing, aggression, and co-occurring problems was associated with lower PGS for general cognitive ability (IQ; β = –0.05 to –0.09, FDR < 0.05) and educational attainment (β = –0.05 to –0.10, FDR < 0.05). Additionally, higher PGS for PTSD was associated with increased risk of internalizing problems at both ages (three years: β = 0.03, p = 0.01, FDR = 0.048; five years: β = 0.05, p = 0.01, FDR = 0.038).

We also observed an age-dependent association between genetic risk of ADHD and behavioral problems: at age three, higher ADHD PGS was linked to greater aggression, but at age five, it was associated with higher internalizing tendencies. A similar pattern was found for depression and neuroticism (PGS-NEU). Notably, PGS-NEU had the broadest associations, significantly correlating with all childhood behavioral problems except for the aggression vs. internalizing contrast (FDR < 0.05).

For brain structure, only the PGS for pallidum volume was associated with co-occurring problems at age five (left: β = 0.16, p = 1.73 × 10⁻⁴, FDR = 0.034; right: β = 0.18, p = 4.51 × 10⁻⁵, FDR = 0.018).

Overall, internalizing problems were associated with the widest range of genetic risk factors, followed by aggression, and then co-occurring problems, suggesting partly distinct genetic architectures underpinning each phenotype. Our negative control analysis (hair-color PGS) showed no significant correlations, further supporting the robustness of these findings (**Suppl. Tables S5** and **S6**, **Suppl. Figs. S5-S13**).

### DNA Methylation and Co-Occurring Behavioral Problems

#### Epigenome-Wide Association Studies (EWAS)

We performed EWAS on 407,705 CpG sites, analyzing DNAm data from up to 3,958 children (**Fig. 1B**). Given the influence of maternal behaviors during pregnancy on fetal DNAm^31–33^, we adjusted for maternal smoking, gestational age, and delivery mode (see **Methods**).

No significant DMPs were identified for internalizing and aggression. However, for co-occurring problems, we identified 101 significant DMPs (p < 10⁻⁷) (five at age three, 96 at age five) (**Fig. 3A**, **Table 1**, **Suppl. Figs. S14-S19** and **Suppl. Tables S7-S8**). Notably, cg13650260 (chr9:129,642,311, hg19) was associated with co-occurring problems at both ages (three years: β = –0.02, p = 1.89 × 10⁻⁸; five years: β = –0.02, p = 3.05 × 10⁻¹⁰). The closest gene, ZBTB34, has been linked to schizophrenia, depression, educational attainment, cortical thickness, and immune-related traits^16,34–36^. We did not observe inflated statistics due to cryptic relatedness nor unaccounted confounding factors (lambda: 0.82-0.98; **Methods**; **Fig. 2B** and **2D**). In addition, the Volcano plots for the effects of methylations did not show obvious abnormalities (**suppl. Figs. S16** and **S19**).

**Figure 3.**
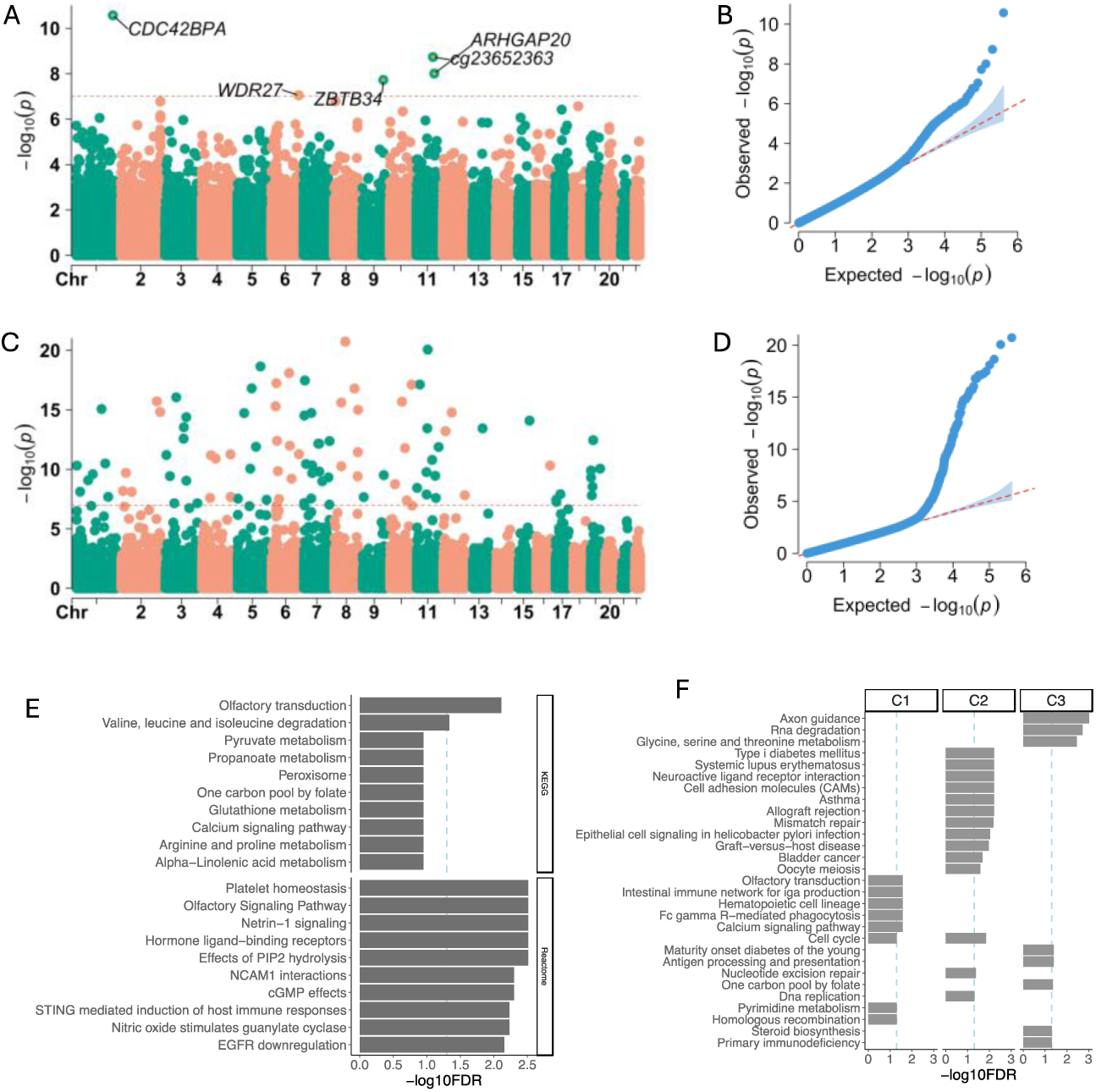
The effect of DNA methylation levels on child behavioral problems. A, B, Manhattan plot (A) and corresponding QQ plot (B) for the EWAS of DNA methylation effects on co-occurring behavioral problems measured at age 3 years. C,D, Manhattan plot (C) and corresponding QQ plot (D) for the EWAS of DNA methylation effects on co-occurring behavioral problems measured at age 5 years. E, Gene sets from the KEGG and Reactome databases enriched for genes identified in A and C.F, KEGG gene sets enriched in gene clusters derived from genes identified in A and C. Closest genes to significant CpG sites are shown. Dotted and dashed lines in A, C, E, and F indicate genome-wide significance thresholds after multiple testing correction.

#### Gene Set Enrichment Analysis (GSEA)

To improve the biological interpretation of the implicated genes in co-occurring problems, we performed gene set enrichment analysis using GeneNetwork v2, a tool based on RNA co-expression networks from 34,119 multi-tissue samples^37^. We combined genes identified from differentially methylated positions (DMPs) at both ages for enrichment analysis. Using two curated databases, we identified two significantly enriched gene sets from Kyoto Encyclopedia of Genes and Genomes (KEGG) ^38^ and 36 from Reactome^39^ at FDR ≤ 0.05 (**Suppl. Tables S9** and **S10**). Figure 3E presents the top ten enriched gene sets from each database. The identified gene sets are primarily associated with neurodevelopment, amino acid metabolism, and immune function (**Fig. 3E** and **Suppl. Tables S9** and **S10**). Notable pathways include axon guidance, olfactory transduction/signaling, and NCAM1 interaction.

In addition to direct gene set enrichment, we applied the GeneNetwork clustering model to group genes associated with co-occurring behavioral problems from EWAS into functional clusters. Three gene clusters were obtained (**Fig. 3F**, **Suppl. Fig. S20** and **Table S11**). Due to the functional homogeneity of clustered genes, we identified more enriched gene sets than in the unclustered analysis, with 29 gene sets from KEGG and 146 from Reactome (**Fig. 3F**, **Suppl. Fig. S21**). KEGG enrichment included additional neurodevelopmental pathways, such as neuroactive ligand-receptor interaction, axon guidance, and calcium signaling. Several immune system-related pathways were also significantly enriched. Similarly, Reactome enrichment highlighted multiple developmental and immune-related gene sets (**Suppl. Fig. S21**).

#### Co-Methylation Modules for Co-Occurring Behavioral Problems

Since co-occurring behavioral problems showed the highest variance explained by DNA methylation and yielded a large number of EWAS signals, we performed co-methylation module analysis using WGCNA^40^ separately for ages three and five (**Methods**). We identified seven co-methylation modules at age three and eight modules at age five (**Figs. 4A** and **4B**). At both ages, the black module eigengenes explained the largest proportion of module variance (age three: 41%, age five: 51%) (**Suppl. Tables S12** and **S13**). To examine their association with behavioral outcomes, we tested the eigengenes for each module against co-occurring behavioral problems. At age three, the black module eigengene was nominally associated with co-occurring behavioral problems (p = 1.44 × 10⁻⁴), with lower eigengene levels linked to increased risk of co-occurring behavior problems (**Figs. 4C** and **4D**; **Suppl. Tables S14** and **S15**).

**Figure 4.**
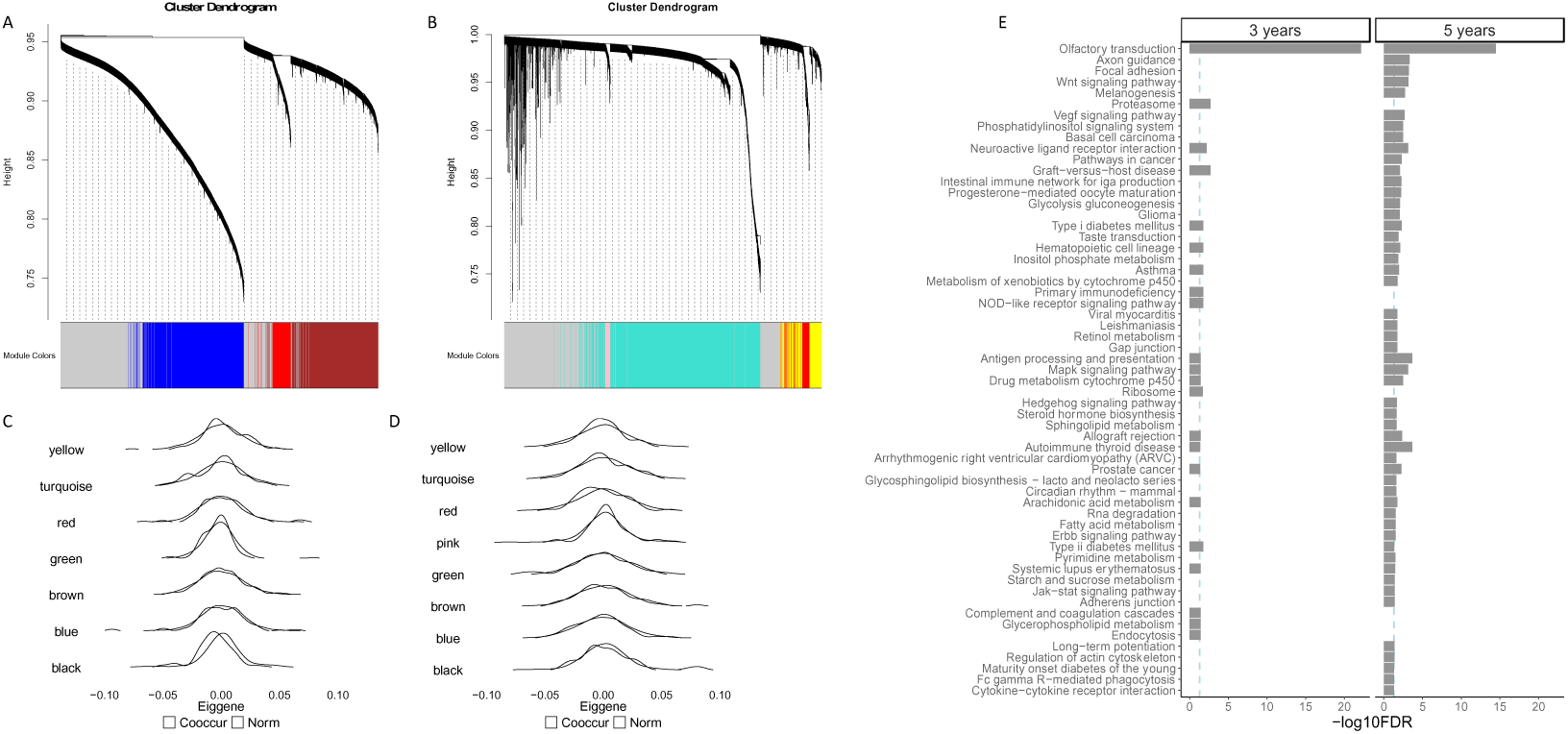
Co-methylation modules for the cooccurring behavioral problem. A,B, Dendrograms for the co-methylation modules associated with co-occurring behavioral problems at age 3 (A) and age 5 (B). Colors indicate distinct modules. C,D, Associations between the eigengenes of the top-ranked modules (black) and co-occurring behavior at age 3 (C) and age 5 (D). Filled colors represent behavioral groups. E, Gene set enrichment analysis from the KEGG database for genes belonging to the black modules identified at ages 3 and 5. Dashed lines indicate statistical significance thresholds after multiple testing correction.

We further performed GeneNetwork analysis on genes assigned to the black module at both ages (**Fig. 4E**). Consistent with EWAS results, more enriched gene sets were identified at age five than at age three. Several gene sets were enriched at both ages, including olfactory transduction and neuroactive ligand-receptor interaction. Notably, axon guidance was enriched at age five but not at age three, suggesting potential age-specific methylation effects. In addition to neurodevelopmental processes, several immune– and metabolism-related gene sets were identified at only one age, reinforcing the hypothesis that multiple biological processes contribute to co-occurring behavioral problems beyond neurodevelopment alone (**Fig. 4E**; **Suppl. Tables S16** and **S17**). Reactome enrichment yielded 91 gene sets at age three and 94 at age five, further supporting the involvement of developmental, immune, and metabolic pathways in childhood behavioral risk (**Suppl. Figs. S21** and **S22**, **Tables S18** and **S19**).

#### Causal Effects of DNA Methylation on Co-Occurring Problems

To determine whether DNAm changes causally influence co-occurring behavioral problems, we performed cis-mQTL analysis on the 101 DMPs identified in EWAS and applied the Mendelian randomization (MR) framework. We found mQTLs for 26 of these CpG sites (**Fig. 5A** and **Suppl. Tables S20-S45**). Using MR, we tested whether DNAm at these CpG sites had a causal effect on co-occurring problems. We identified one nominally significant association: higher methylation levels at cg06772671 increased the risk of co-occurring problems at age five (MR p = 0.037, Heidi p = 0.45) (**Suppl. Table S46**). The closest gene, KIAA0513, has been linked to synaptic function, apoptosis, and neuroplasticity^41^.

**Figure 5.**
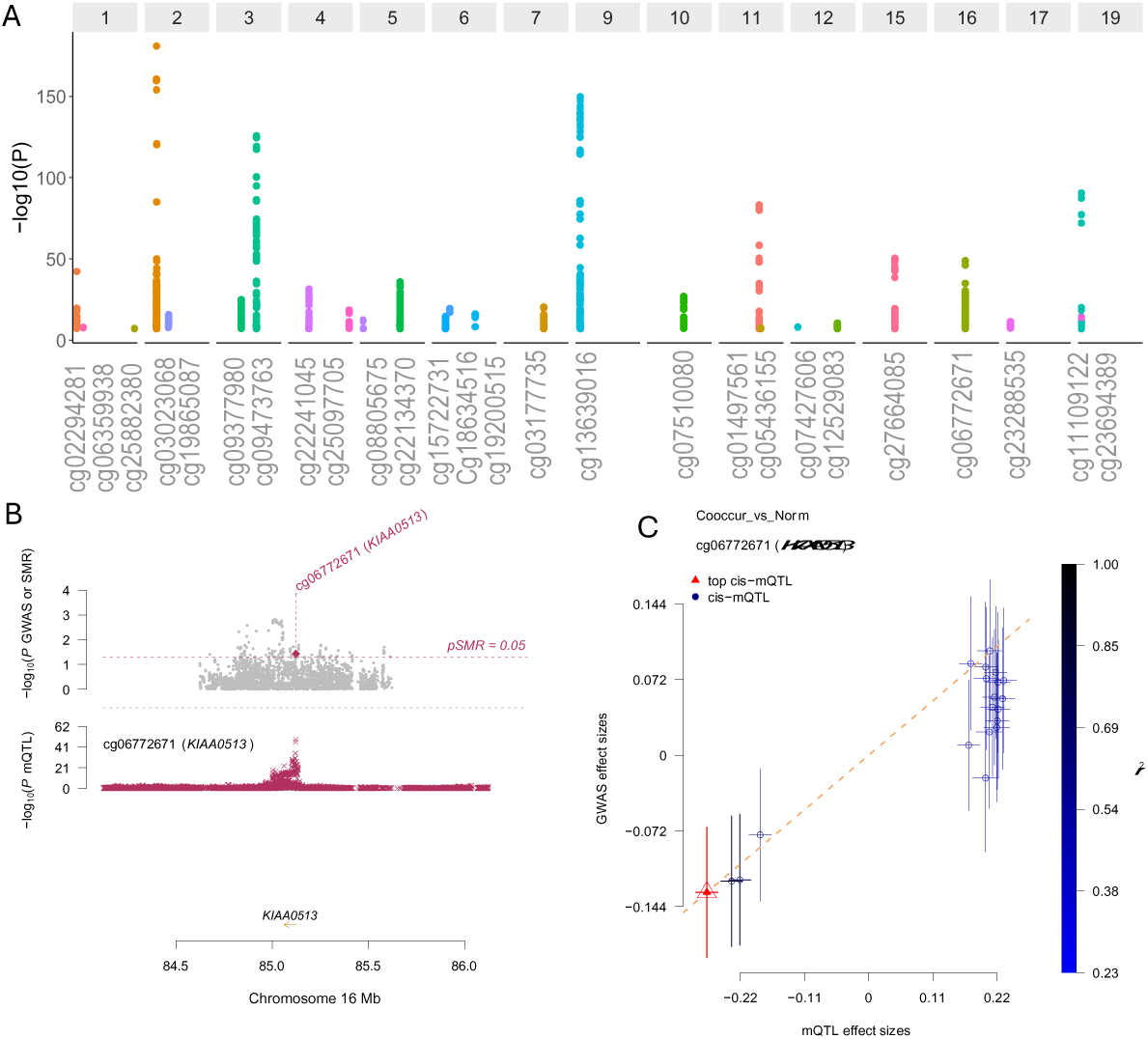
mQTL and Mendelian randomization analysis on DNA methylation sites associated with the cooccurring behavioral problem. A, mQTL associations for CpG sites identified by EWAS for the co-occurring behavioral problem. CpG identifiers are shown on the x-axis, and association strengths (−log₁₀ p-values) are shown on the y-axis. Chromosome numbers are indicated in the panel headers. B, Associations between the cis-SNP, methylation levels at cg06772671, and co-occurring behavior. C, Mendelian randomization estimates of the causal effect of methylation at cg06772671 on the risk of co-occurring behavior.

## DISCUSSIONS

Our findings provide genetic and epigenetic evidence that co-occurring internalizing and externalizing behavioral problems in childhood has a unique biological underpinning. Using a person-centered approach, we identified distinct behavioral profiles marked by these co-occurring symptoms, highlighting meaningful subgroups that may follow different developmental and biological pathways. Importantly, the relative contributions of genetic and epigenetic factors appear to shift with age, underscoring the dynamic nature of behavioral development.

Our DNAm analyses suggests that epigenetic regulation plays a role in childhood co-occurring problems, particularly at age five, whereas internalizing and aggression problems did not show significant DNAm associations. Since our DNAm data were derived from umbilical cord blood, only prenatal influences—either genetic or environmental—could shape these DNAm levels. Our mQTL analysis identified a large number of genetic variants associated with DNAm. Yet Mendelian randomization provided little evidence that DNAm mediates genetic effects on behavior. This suggests that observed DNAm differences may be driven by prenatal environmental factors rather than genetic predisposition. While we adjusted for maternal smoking, gestational age, and delivery mode—known contributors to fetal DNAm variation— we cannot rule out the possibility that other, unmeasured prenatal environmental exposures may have influenced both DNAm patterns and co-occurring behavioral problems through parallel pathways. These findings align with recent EWAS studies in adult populations, where causal DNAm effects on behavior remain difficult to establish ^42^. However, our ability to detect causal relationships may have been limited by the relatively small sample size for instrumental variable analyses.

Our GWAS results identified the *USP12* locus, significantly associated with co-occurring problems. This has previously been linked to risk-taking behavior in adults^30^. This is particularly interesting in light of prior evidence linking childhood risk-taking behaviors to persistent co-occurring problems later in life^43^, suggesting developmental continuity between these traits. Our results confirm that common genetic variants contribute to childhood behavioral variation, with SNP-heritability estimates comparable to those observed for adult behavioral traits and lifestyle factors but lower than those for psychiatric disorders ^44–46^. The highly polygenic nature of child behavioral problems has been underexplored in prior studies.

One particularly intriguing finding is that high PGS for ADHD was associated with both internalizing and externalizing problems at different ages. This suggests that ADHD might be more closely related to co-occurring behavioral problems rather than being confined to a strictly externalizing phenotype. Moreover, the relationship between PGS-ADHD and aggression versus internalizing behavior shifted between ages three and five: higher ADHD genetic risk was associated with greater aggression at age three but with internalizing behavior at age five. These findings align well with previous reports^47–49^. One possible explanation could be that at age three, ADHD genetic risk may be driven by impaired inhibitory control and heightened impulsivity, and thus primarily manifest as aggression. However, by age five, as cognitive and emotional regulation processes mature, the same genetic predisposition may shift toward internalizing difficulties, such as anxiety and emotional dysregulation. While the biological basis of this shift remains unclear, recent findings suggest that developmental changes in symptom expression can be traced through repeated behavioral assessments within individuals^21^. Our polygenic score analyses align with findings from genomic structural equation modeling^50^, reinforcing the association between PGS for depression and PTSD with internalizing problems. However, methodological differences in how latent factors are extracted preclude direct comparisons.

Further distinguishing co-occurring problems from internalizing or externalizing behaviors is their unique genetic association with pallidum volume, a subcortical structure involved in cognition and movement^51^. This may suggest that distinct neurodevelopmental mechanisms may underlie the persistence and severity of co-occurring behavioral problems, warranting further investigation into brain structural correlates of early behavioral traits.

To gain insight into potential biological pathways, we performed gene set enrichment analyses using EWAS-derived genes and co-methylation modules. Despite differences in methodology, both approaches highlighted neurodevelopmental, immune-related, and metabolic pathways. These findings suggest that prenatal factors influencing these biological processes may have long-term effects on child behavior, persisting at least into early childhood.

Several aspects strengthen our study. The large sample size of children with both genetic and DNA methylation (DNAm) data enabled us to distinguish genetic versus environmental influences on childhood behavior. The use of umbilical cord blood DNAm profiling minimized the confounding effects of postnatal environmental factors, which may explain the strong DNAm signals observed in our EWAS analysis. Additionally, the longitudinal assessment of child behavior at ages three and five provided a unique opportunity to examine age-related shifts in genetic and epigenetic contributions. Future research incorporating multi-omics approaches and longer follow-ups will be crucial in validating and expanding our findings.

Our study has some limitations. The categorization of child behavior into four discrete groups may have oversimplified the dimensional nature of behavioral traits^8^. While our person-centered approach provides valuable insights by comparing co-occurring behavioral profiles to a normative group, it’s important to recognize that these profiles are specific to our dataset and may not be generalizable to other samples. Additionally, the classify-analyze method we used does not account for uncertainty in profile membership, which could lead to biased estimates or understated variability when comparing the co-occurring and normative groups^52^. Although the use of consistent questionnaires across ages ensured comparability, a more comprehensive longitudinal design would enable deeper insights into the progression of behavioral patterns over time. Finally, the absence of an independent replication sample highlights the need for large-scale studies to validate our findings.

In conclusion, our study demonstrates that co-occurring childhood behavioral problems have different genetic and biological features compared to “pure” internalizing or externalizing behaviors. The co-occurrence of internalizing and externalizing problems presents particular challenges for diagnosis and intervention, as children with comorbid symptoms often follow more severe and persistent developmental trajectories, than those with isolated symptoms. By integrating genetic, epigenetic, and pathway analyses, we provide new insights into age-dependent genetic and DNAm influences on co-occurring behavioral problems. These findings highlight the importance of considering developmental changes in behavioral genetics research and suggest that early-life interventions targeting co-occurring problems may help mitigate long-term mental health risks. Future studies should continue to explore early risk factors and causal pathways, with the goal of informing targeted prevention and intervention strategies in early childhood.

## METHODS

### Sample and data processing

The MoBa (Mother, Father, and Child Cohort) study is a Norwegian pregnancy cohort initiated in 1999. By the conclusion of data collection in 2008, the cohort included 95,200 mothers, 114,500 children, and 75,200 fathers^28^. The present study focuses on child data, with umbilical cord blood samples collected at birth for most participants. This study is based on version 12 of the quality-assured MoBa dataset, released for research in 2021. The establishment of MoBa and initial data collection were approved by the Norwegian Data Protection Agency and the Regional Committees for Medical and Health Research Ethics (REK). The MoBa cohort is currently regulated by the Norwegian Health Registry Act. This study received ethical approval from REK (2009/1899-7; 2013/2061) and the Norwegian Data Inspectorate, with informed consent obtained from all participants prior to participation.

Child behavior was assessed using the Child Behavior Checklist (CBCL^53^), as described in Tsotsi et al. (2023). Mothers rated their children’s behaviors on a three-point Likert scale ranging from 0 (not true) to 2 (very true). Internalizing difficulties were measured using nine items at age three and ten items at age five, with raw scores ranging from 0 to 18 at age three and 0 to 20 at age five. Aggression was assessed using four items at both time points, with raw scores ranging from 0 to 8.

Latent profile analysis was performed separately for ages three and five to identify behavioral subgroups. Based on model performance, four groups were identified at each age (**Suppl Figs S23 and S24**): a group with low or normal levels of behavioral problems (Norm), a group with primarily internalizing difficulties (Internal), a group with primarily aggressive behaviors (Aggres), and a group with co-occurring internalizing and aggressive behaviors (Cooccur). The distribution of children across these groups at each age is shown in **Figure 1B**.

### Genotypes and Imputation

The MoBa genetics data is managed by a centralized core team, with strict quality control procedures applied before data sharing across subprojects. The present study is based on version v1 of the dataset, which was generated using a carefully designed pipeline that accounts for the unique family structure in the MoBa study while performing imputation^54^. This pipeline imputed and released 6,981,748 hard-coded autosomal SNPs for 207,569 unique individuals of European ancestry, along with sex chromosome genotypes for a subset of individuals. The present study did not use sex chromosome data or parental genotypes. Among the children with available behavioral data, up to 40,149 have quality-controlled genotype data (**Fig. 1B**), though the exact number varies by behavioral and age group.

Genotype-based principal components (PCs) were computed following standard procedures. Relatively rare SNPs, defined as those with a minor allele frequency (MAF) below 0.05, and SNPs failing Hardy-Weinberg equilibrium tests (p < 10⁻⁶) were removed. The remaining SNPs were thinned using a sliding window approach to remove highly correlated neighboring SNPs, as implemented in PLINK 2^55^ with the parameters –indep-pairwise 100 50 0.1. The resulting thinned genotype data were then used as input for principal component analysis (PCA) using the PLINK –pca command. The top ten PCs were included in downstream analyses to account for subtle population stratification.

### DNA Methylation

DNA methylation profiles for MoBa participants were processed and quality-checked by a core team across ten data waves, following the pipeline documented at https://github.com/folkehelseinstituttet/mobagen/wiki/Methylation#met001. Briefly, preprocessing was conducted separately for each wave using the minfi R package^56^, following the 12-step protocol outlined in the pipeline.

The present study includes only child data and excludes waves designed for specific drug effects (waves 006 and 007, focused on Paracetamol) or those that included only ADHD patients. Waves 001 to 003 were based on the Illumina Infinium 450K BeadChip (Illumina, CA), while the remaining waves used the Illumina Infinium EPIC BeadChip (Illumina, CA). The quality-checked data were merged into a single dataset, retaining only CpG sites present on both array types. To mitigate batch effects, both wave indicators and chip type were included as covariates in all downstream analyses. As with genetic data, the number of unique individuals varies across behavioral groups (**Fig. 1B**).

### Statistical Analysis

#### Genome-Wide Association Studies (GWAS)

Given the sample size constraints relative to previous GWAS studies, we employed a case-control design instead of analyzing raw behavioral scores as continuous traits. We performed GWAS for four behavioral contrasts: internalizing versus normal, aggression versus normal, co-occurring versus normal, and aggression versus internalizing. This approach, to some extent, mimics the classic extreme traits design, which can enhance power for gene discovery, though it may lead to upwardly biased effect size estimates.

Approximately 5% of the included children are siblings. To account for family structure, we used generalized linear mixed-effects models, where family identifiers were included as random intercept effects. Fixed effects included the SNP, the top ten principal components (PCs), sex, and genotyping batch indicators. The genetic relationship matrix was computed using the same thinned genotype data as for the PC analysis. GWAS was performed using GLMM models implemented in GCTA v1.94^57^ with the command –fastGWA-mlm-binary. Genome-wide significance was defined as p < 5 × 10⁻⁸.

#### SNP heritability

The proportion of phenotypic variance attributable to genome-wide SNPs was estimated using GCTA’s restricted maximum likelihood models. The significance of SNP-heritability estimates was assessed using the likelihood ratio test implemented in GCTA with the –reml command. To correct for multiple testing, we applied the Benjamini-Hochberg false discovery rate (FDR), considering FDR ≤ 0.05 as statistically significant.

#### Polygenic Risk Score (PGS) Analysis

To investigate genetic correlations between childhood behavioral problems and major neuropsychiatric conditions, brain structural variations, and socioeconomic traits, we conducted a PGS analysis. Summary statistics were obtained from GWAS on schizophrenia (SCZ)^16^, bipolar disorder (BIP)^15^, depression (DEP)^58^, autism spectrum disorder (ASD)^59^, attention deficit hyperactivity disorder (ADHD)^60^, post-traumatic stress disorder (PTSD)^61^, generalized anxiety disorder (ANX)^62^, drinks per week (DRK)^63^, cigarettes smoked per day (SMK)^63^, general intelligence (IQ)^64^, educational attainment (EDU)^65^, neuroticism (NEU)^66^, and obsessive-compulsive disorder (OCD)^67^. We also included 101 GWAS-derived brain structural traits^68^. As a negative control, we used hair color (HC)^69^. Summary statistics were aligned to high-quality SNPs from the HapMap3 dataset before analysis. To account for winner’s curse bias in SNP effect sizes, we applied a Bayesian mixture model, PRS-CS^70^. The posterior-corrected SNP effects were used as allele weights in computing PGS via the PLINK –score function.

We used multiple logistic regression models to assess the effect of polygenic scores (PGS) on the probability of exhibiting abnormal behavior compared to the normal group. The same covariates used in the GWAS analysis were included in the PGS models. To correct for multiple testing, we applied the false discovery rate (FDR) procedure separately for brain structure PGS and the remaining PGS analyses. Results were considered statistically significant at FDR ≤ 0.05.

#### Epigenome-Wide Association Studies (EWAS)

Based on the merged DNA methylation dataset, we performed EWAS using the MOMENT model^71^, which, despite its conservative nature, effectively reduces batch effects by explicitly modeling neighboring methylation sites. Because DNA methylation levels can be either causal or consequential, this study tested whether methylation levels influence the probability of being in an abnormal behavioral group compared to the normal group.

In total, we analyzed 407,705 cytosine-phosphate-guanine (CpG) sites. To minimize confounding, we included gestational age, maternal smoking during pregnancy, delivery mode, profiling wave indicator, and BeadChip type indicator as covariates. We applied a p-value threshold of ≤ 1 × 10⁻⁷ to identify differentially methylated positions (DMPs).

#### Variance in Behavioral Traits Explained by DNA Methylation

We estimated the proportion of variance in behavioral traits attributable to genome-wide DNA methylation variation using OSCA software. The same set of covariates used in the EWAS analysis was included in these models. Multiple testing correction was performed using the Benjamini-Hochberg FDR procedure, with FDR ≤ 0.05 considered statistically significant.

#### DNA Methylation Quantitative Trait Locus (mQTL) Analysis

For each identified DMP, we conducted cis-mQTL analysis using GCTA with the –fastGWA-mlm command. SNPs located within a 1 Mb radius of a DMP were analyzed. The same covariates used in the GWAS analysis were included in the models to control for confounding effects. To correct for multiple testing, we applied the Bonferroni correction.

#### Mendelian Randomization (MR) Analysis

We performed Mendelian randomization (MR) analysis to test whether DNA methylation levels are causally associated with childhood behavioral problems. To increase statistical power and reduce bias, we focused only on traits with significant EWAS signals and DMPs that had significant mQTLs. Instrumental SNPs were selected based on the EWAS significance threshold, with highly correlated SNPs (r² > 0.1) removed to ensure independent instruments.

MR analysis was conducted using SMR software ^72^, which is specifically designed for molecular traits as exposures. The HEIDI test implemented in SMR was used to detect horizontal pleiotropy, with HEIDI p < 0.05 indicating potential pleiotropic effects. Evidence for a potential causal relationship between DMPs and childhood behavioral problems was defined as SMR p ≤ 0.05 and HEIDI p > 0.05.

#### Weighted Correlation Network Analysis (WGCNA)

To aid in the biological interpretation of EWAS findings, we applied weighted correlation network analysis (WGCNA)^40^ to genes identified by EWAS. This analysis was restricted to behavioral traits for which significant EWAS signals were detected. For these traits, CpG sites with nominal significance in EWAS (p ≤ 0.05) were selected, and Beta-values were converted to M-values to better fit the WGCNA model assumptions.

We used the pickSoftThreshold function in the WGCNA package to determine the optimal power parameter for the scale-free network assumption, scanning values from 1 to 20 (**Suppl. Figs. S23** and **S24**). The selected power, along with M-values of the identified CpG sites, was used as input for blockwiseModules, with default settings for all other parameters. The identified co-methylation networks were assigned unique colors for reference. Genes located near CpG sites were assigned to the corresponding modules. To summarize the functional relevance of these modules, eigengenes (the first principal component of the co-methylation networks) were computed using moduleEigengenes. The association between eigengenes and behavioral groups was assessed using logistic regression, adjusting for the same covariates used in EWAS to control for confounding.

#### Gene Set Enrichment Analysis

To infer the biological functions of the identified genes, we performed gene set enrichment analysis using the online tool GeneNetwork v2^37^ (https://www.genenetwork.nl/). We first set the average prioritization z-score threshold to 2.07 and then performed enrichment analysis for two databases: KEGG^38^ and Reactome^39^. Since many gene sets within each database share genes and are therefore correlated, we applied the FDR procedure to correct for multiple testing, considering FDR ≤ 0.05 as the significance threshold. GeneNetwork analysis was conducted separately for genes identified from EWAS and genes associated with the most significant WGCNA modules. Wherever possible, analyses were also performed separately for data from ages three and five. GeneNetwork further allows the construction of gene clusters based on RNA co-expression data from 31,449 multi-tissue samples. We derived such clusters separately for genes identified by EWAS and WGCNA modules to explore potential functional relationships.

## Supporting information

Supplementary Information is in Suppl_final.doc

## Acknowledgements

We are grateful to all the participating families in Norway who take part in this on-going cohort study. For generating high-quality genomic data, we thank the Norwegian Institute of Public Health (NIPH), the HARVEST collaboration, the NORMENT Centre at the University of Oslo, the Center for Diabetes Research at the University of Bergen, deCODE Genetics, the Research Council of Norway, the South-Eastern and Western Norway Regional Health Authorities, the ERC AdG, Stiftelsen KG Jebsen, the Trond Mohn Foundation, and the Novo Nordisk Foundation. **Study funding:** The Norwegian Mother, Father and Child Cohort study is supported by the Norwegian Ministry of Health and the Norwegian Research Council/FUGE (grant number 151918/S10). This work was also supported by the Research Council of Norway grant number 288083 and 301004 and by the Research Council of Norway FRIPRO (no. 302854). In addition, LJH is supported by funding from the Norwegian South-East Regional Health Authority (Helse South-East;2022083).

## Data availability

Genome-wide summary statistics for child behavior problems are available at http://figshare/XXXX. GWAS summary statistics for psychiatric disorders (SCZ, BIP, ASD, ADHD, PTSD, DEP, ANX, and OCD) can be downloaded from https://pgc.unc.edu/for-researchers/download-results/. Summary statistics for drinking and smoking traits are available from https://conservancy.umn.edu/items/, and for IQ and neuroticism from https://cncr.nl/research/summary_statistics/. Data on educational attainment can be found at http://www.thessgac.org/, and brain imaging GWAS results are accessible via https://github.com/BIG-S2/GWAS.

## Code availability

Genome-wide association analysis for child behavior problems was performed using GCTA v1.94 (https://yanglab.westlake.edu.cn/software/gcta/#fastGWA). Polygenic score analysis was conducted with PRS-CS (https://github.com/getian107/PRScs) and PLINK 2.0 (https://www.cog-genomics.org/plink/2.0/). EWAS analysis was performed using the OSCA toolset (https://yanglab.westlake.edu.cn/software/osca). Gene set analysis used the WGCNA R package (https://cran.r-project.org/web/packages/WGCNA/index.html) and https://www.genenetwork.nl/. Mendelian randomization analysis was performed using SMR (https://yanglab.westlake.edu.cn/software/smr/).

