## Supplementary Information is in Suppl_final.doc for "Genetic and Epigenetic Foundations of Childhood Internalizing and Externalizing Problems and their Co-occurrence"

### Fig. S1. The Manhattan plots for GWAS of child behavioural problems measure at age three years.

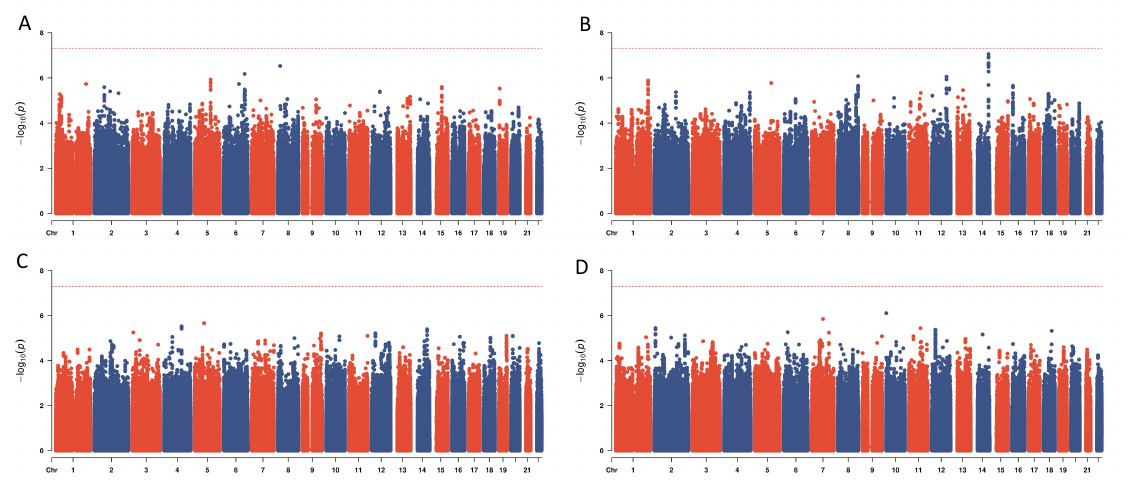

A. Aggressive vs. internalizing behavior; B. aggressive vs. normal behavior; C. co-occurring vs. normal behavior; D. internalizing vs. normal behavior.

### Fig. S2. The QQ plots for GWAS of child behavioural problems measure at age three years.

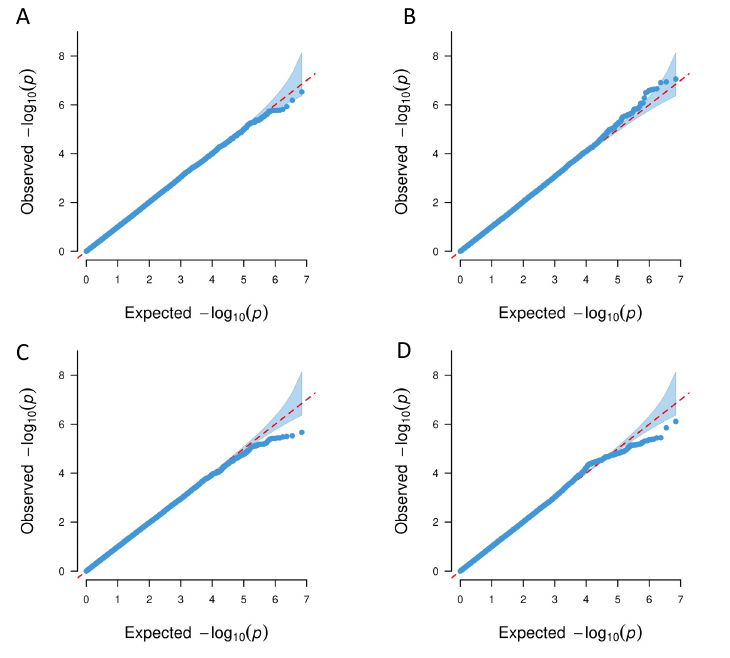

A. Aggressive vs. internalizing behavior; B. aggressive vs. normal behavior; C. co-occurring vs. normal behavior; D. internalizing vs. normal behavior.

### Fig. S3. The Manhattan plots for GWAS of child behavioral problems measure at age five years.

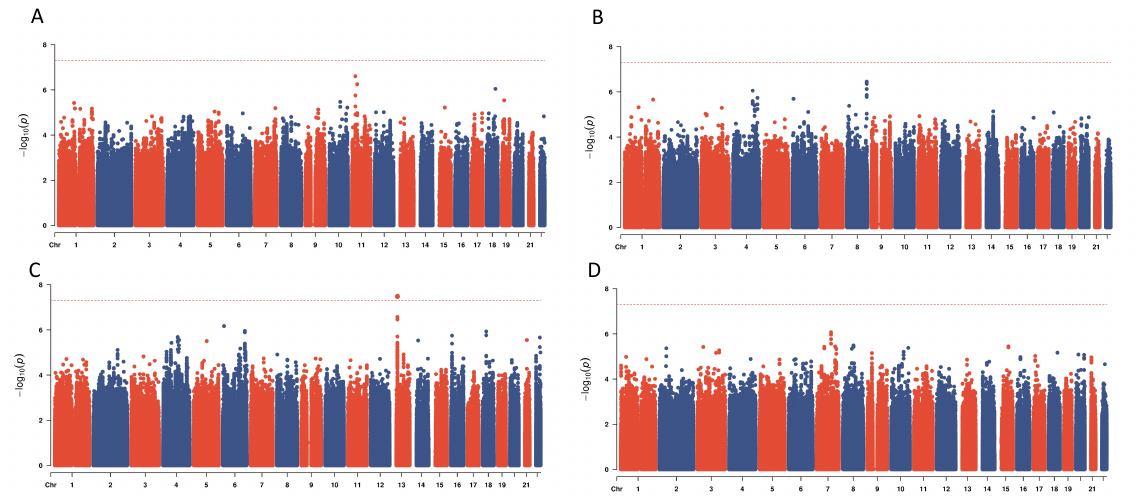

A. Aggressive vs. internalizing behavior; B. aggressive vs. normal behavior; C. co-occurring vs. normal behavior; D. internalizing vs. normal behavior.

### Fig. S4. The QQ plots for GWAS of child behavioural problems measure at age five years.

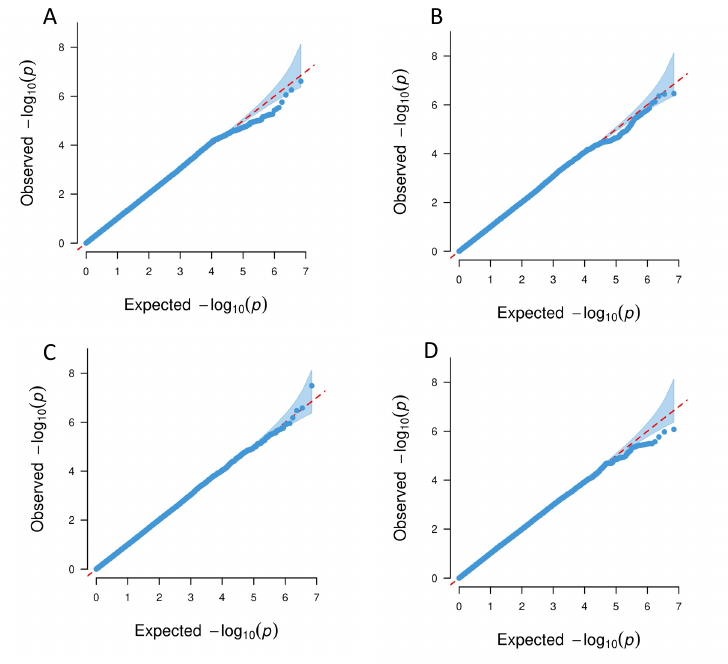

A. Aggressive vs. internalizing behavior; B. aggressive vs. normal behavior; C. co-occurring vs. normal behavior; D. internalizing vs. normal behavior.

### Fig. S5. Distribution of PGS for psychiatric, behavioral and socioeconomic traits for MoBa participants analyzed versus not (missing) at age three years.

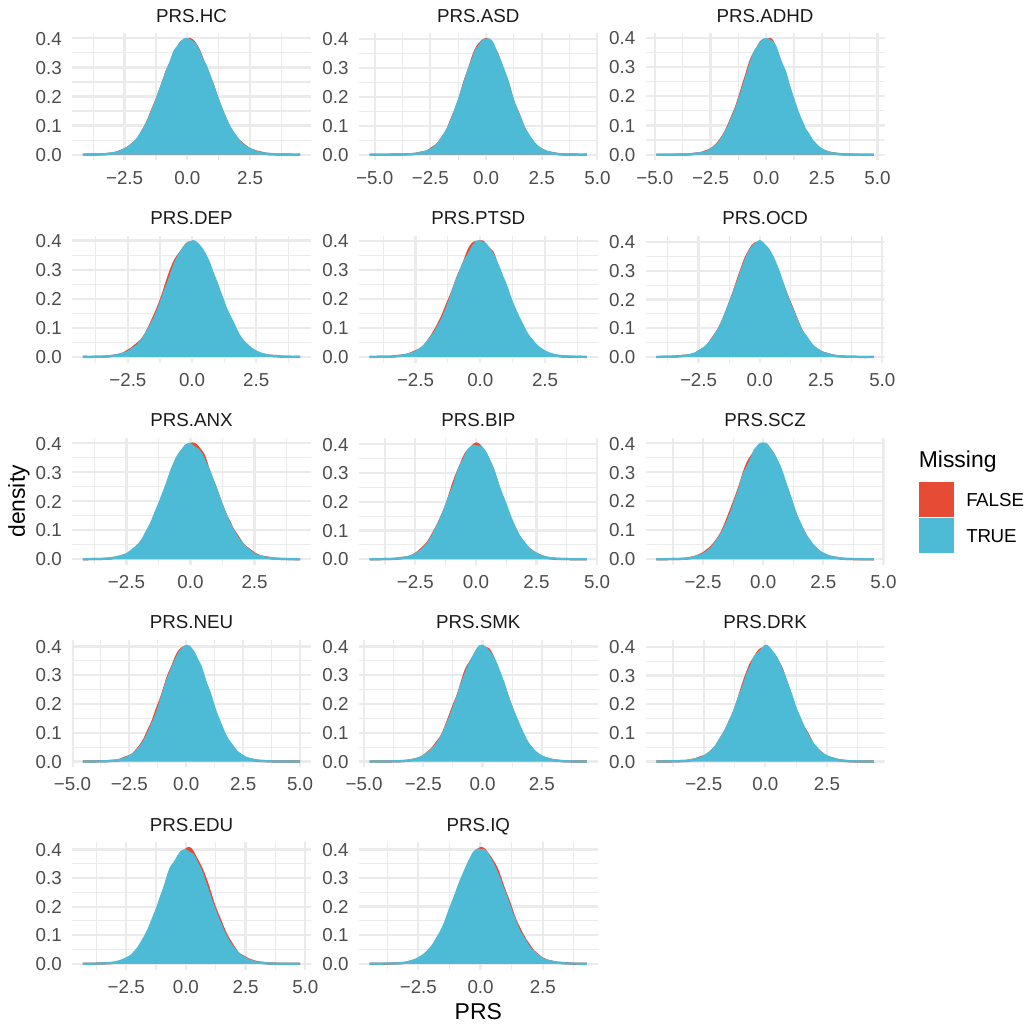

Schizophrenia (SCZ), bipolar disorder (BIP), depression (DEP) , autism spectral disorder (ASD), attention deficit /hyperactivity disorder (ADHD), post-traumatic stress disorder (PTSD), general anxiety (ANX), drinks per week (DRK), smoke per day (SMK), general intelligence (IQ), education attainments (EDU), neuroticism (NEU), Obsessive Compulsive disorder (OCD), and hair color (HC).

### Fig. S6. Distribution of PGS for ROI of brain structural variations for MoBa participants analyzed versus not (missing) at age three years.

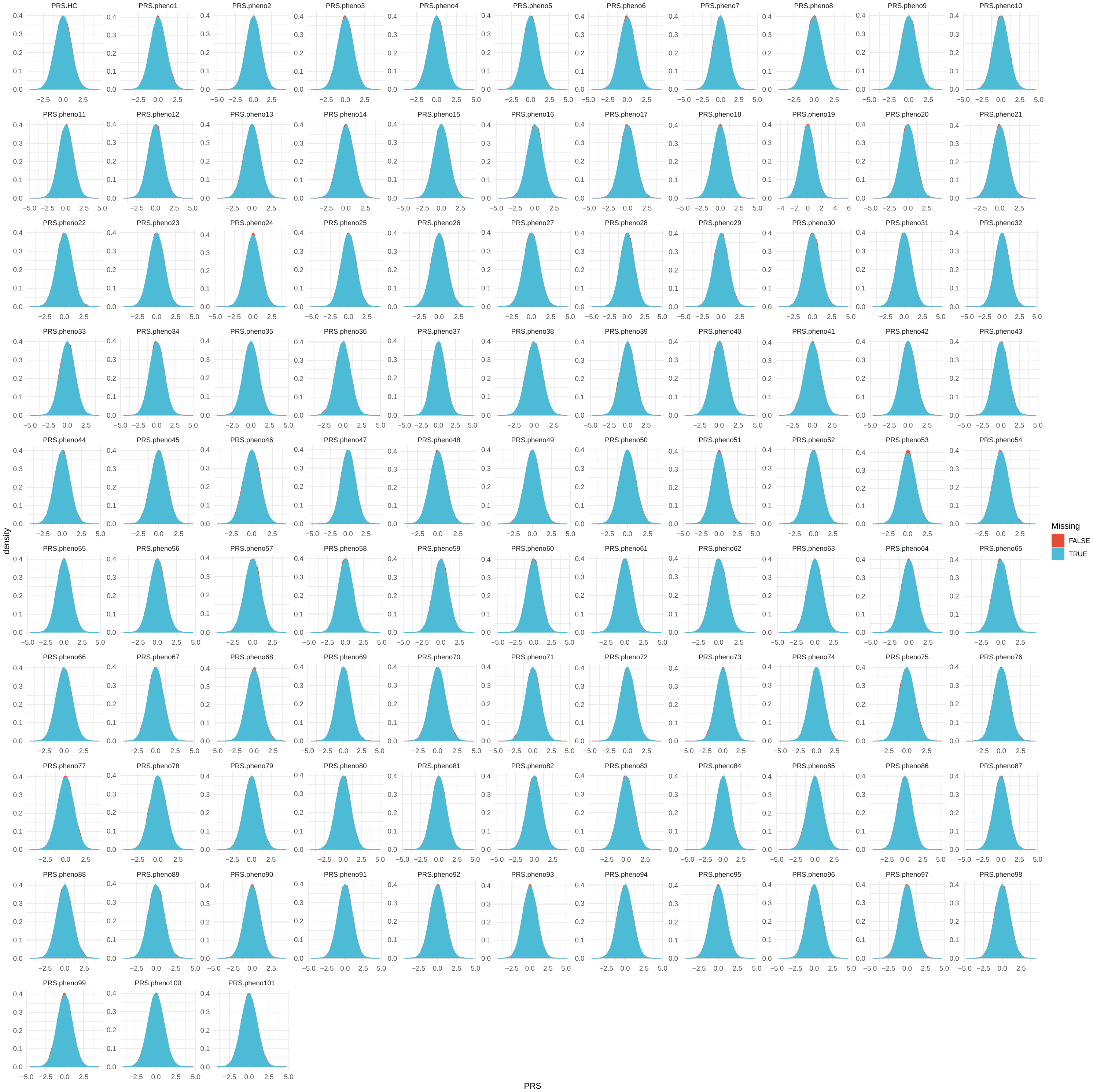

pheno1,left.caudal.anterior.cingulate;pheno2,left.caudal.middle.frontal;pheno3,left.cuneus;pheno4,left.entorhinal;pheno5,left.fusiform;pheno6,left.inferior.parietal;pheno7,left.inferior.temporal;pheno8,left.isthmus.cingulate;pheno9,left.lateral.occipital;pheno10,left.lateral.orbitofrontal;pheno11,left.lingual;pheno12,left.medial.orbitofrontal;pheno13,left.middle.temporal;pheno14,left.parahippocampal;pheno15,left.paracentral;pheno16,left.pars.opercularis;pheno17,left.pars.orbitalis;pheno18,left.pars.triangularis;pheno19,left.pericalcarine;pheno20,left.postcentral;pheno21,left.posterior.cingulate;pheno22,left.precentral;pheno23,left.precuneus;pheno24,left.rostral.anterior.cingulate;pheno25,left.rostral.middle.frontal;pheno26,left.superior.frontal;pheno27,left.superior.parietal;pheno28,left.superior.temporal;pheno29,left.supramarginal;pheno30,left.transverse.temporal;pheno31,left.insula;pheno32,right.caudal.anterior.cingulate;pheno33,right.caudal.middle.frontal;pheno34,right.cuneus;pheno35,right.entorhinal;pheno36,right.fusiform;pheno37,right.inferior.parietal;pheno38,right.inferior.temporal;pheno39,right.isthmus.cingulate;pheno40,right.lateral.occipital;pheno41,right.lateral.orbitofrontal;pheno42,right.lingual;pheno43,right.medial.orbitofrontal;pheno44,right.middle.temporal;pheno45,right.parahippocampal;pheno46,right.paracentral;pheno47,right.pars.opercularis;pheno48,right.pars.orbitalis;pheno49,right.pars.triangularis;pheno50,right.pericalcarine;pheno51,right.postcentral;pheno52,right.posterior.cingulate;pheno53,right.precentral;pheno54,right.precuneus;pheno55,right.rostral.anterior.cingulate;pheno56,right.rostral.middle.frontal;pheno57,right.superior.frontal;pheno58,right.superior.parietal;pheno59,right.superior.temporal;pheno60,right.supramarginal;pheno61,right.transverse.temporal;pheno62,right.insula;pheno63,cerebellar.vermal.lobules.I.V;pheno64,cerebellar.vermal.lobules.VI.VII;pheno65,cerebellar.vermal.lobules.VIII.X;pheno66,left.basal.forebrain;pheno67,right.basal.forebrain;pheno68,Brain.stem;pheno69,CSF;pheno70,X3rd.ventricle;pheno71,X4th.ventricle;pheno72,optic.chiasm;pheno73,left.lateral.ventricle;pheno74,left.inferior.lateral.ventricle;pheno75,left.cerebellum.exterior;pheno76,left.cerebellum.white.matter;pheno77,left.thalamus.proper;pheno78,left.caudate;pheno79,left.putamen;pheno80,left.pallidum;pheno81,left.hippocampus;pheno82,left.amygdala;pheno83,left.accumbens.area;pheno84,left.ventral.DC;pheno85,left.vessel;pheno86,right.lateral.ventricle;pheno87,right.inferior.lateral.ventricle;pheno88,right.cerebellum.exterior;pheno89,right.cerebellum.white.matter;pheno90,right.thalamus.proper;pheno91,right.caudate;pheno92,right.putamen;pheno93,right.pallidum;pheno94,right.hippocampus;pheno95,right.amygdala;pheno96,right.accumbens.area;pheno97,right.ventral.DC;pheno98,right.vessel;pheno99,gray.matter;pheno100,white.matter;pheno101,total.brain.volume;

### Fig. S7. Distribution of PGS for psychiatric, behavioral and socioeconomic traits for MoBa participants analyzed versus not (missing) at age five years.

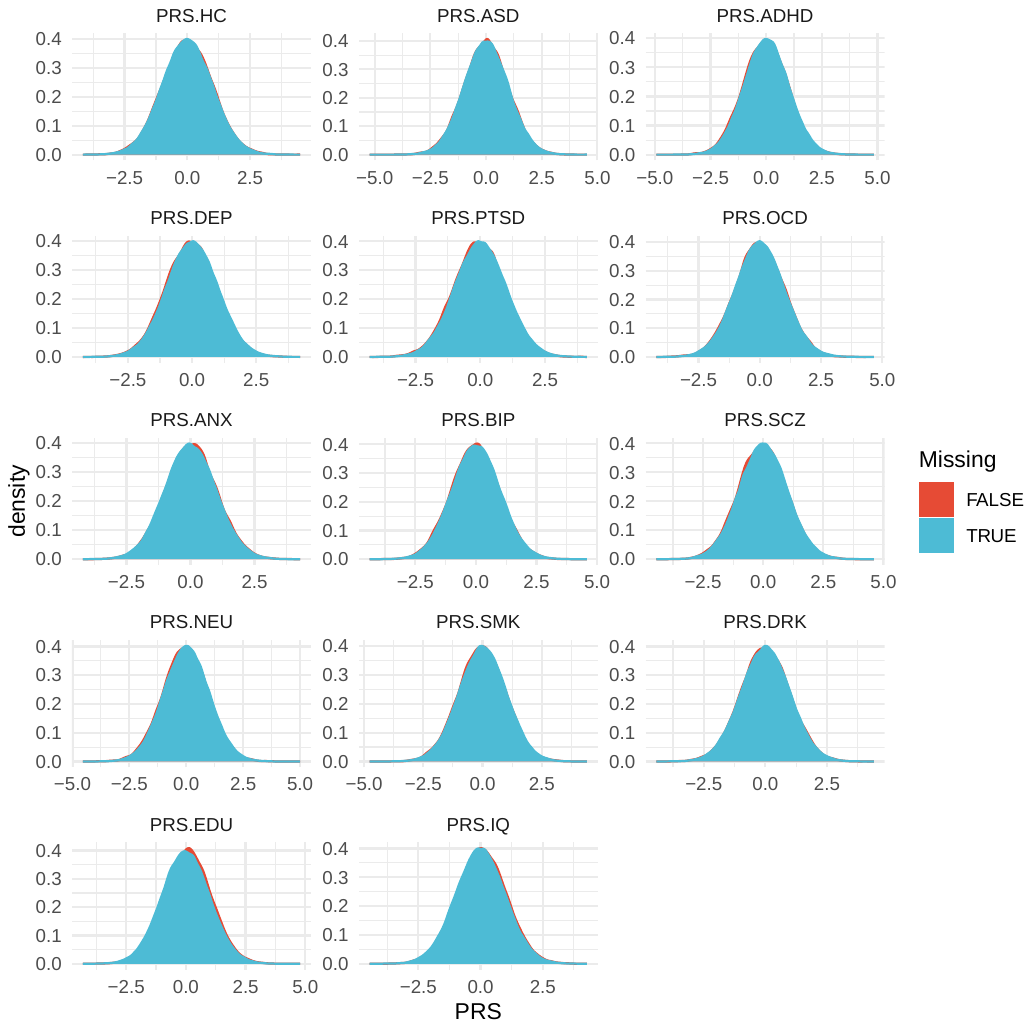

Schizophrenia (SCZ), bipolar disorder (BIP), depression (DEP) , autism spectral disorder (ASD), attention deficit /hyperactivity disorder (ADHD), post-traumatic stress disorder (PTSD), general anxiety (ANX), drinks per week (DRK), smoke per day (SMK), general intelligence (IQ), education attainments (EDU), neuroticism (NEU), Obsessive Compulsive disorder (OCD), and hair color (HC).

### Fig. S8. Distribution of PGS for ROI of brain structural variations for MoBa participants analyzed versus not (missing) at age three years.

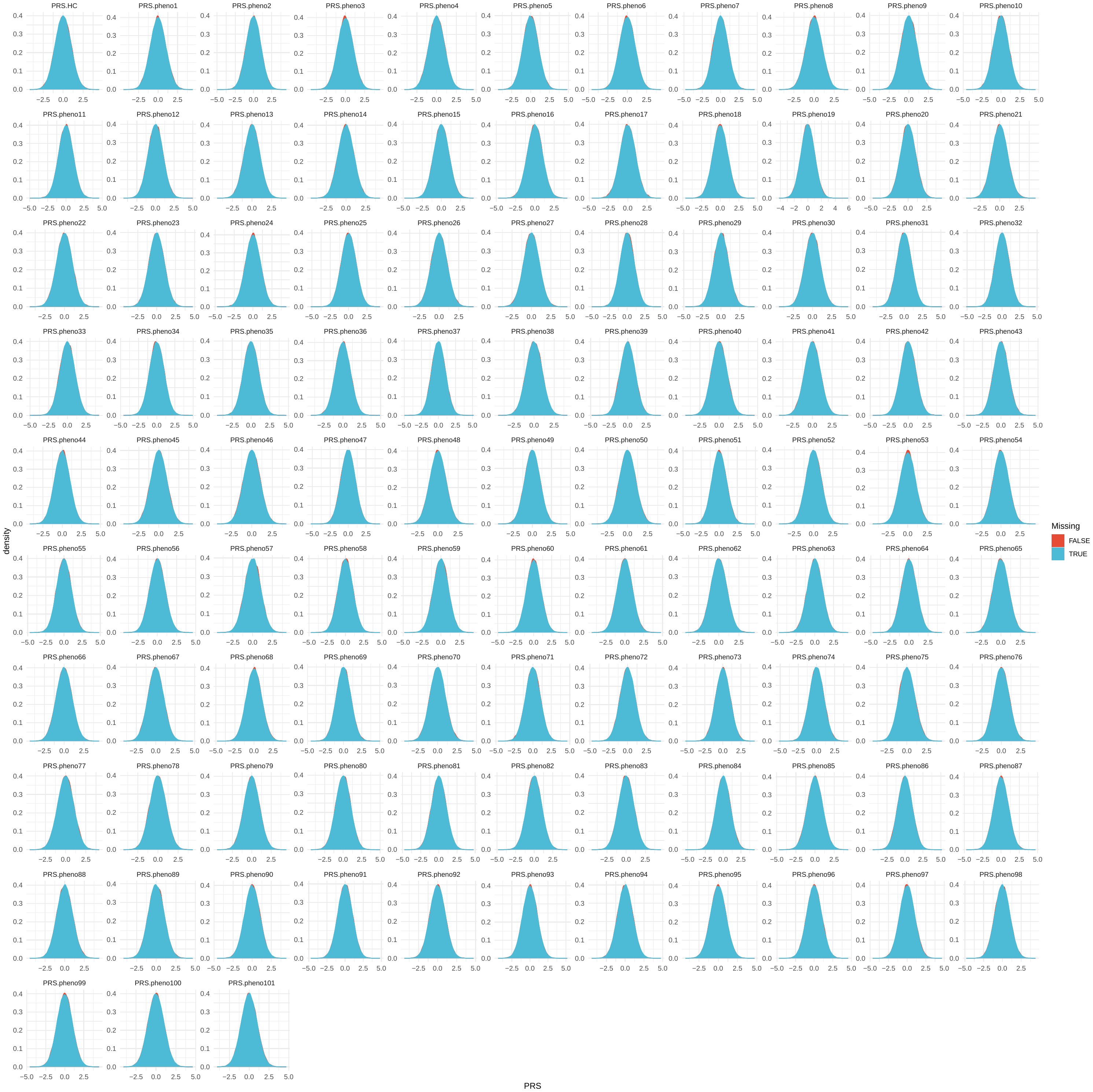

pheno1,left.caudal.anterior.cingulate;pheno2,left.caudal.middle.frontal;pheno3,left.cuneus;pheno4,left.entorhinal;pheno5,left.fusiform;pheno6,left.inferior.parietal;pheno7,left.inferior.temporal;pheno8,left.isthmus.cingulate;pheno9,left.lateral.occipital;pheno10,left.lateral.orbitofrontal;pheno11,left.lingual;pheno12,left.medial.orbitofrontal;pheno13,left.middle.temporal;pheno14,left.parahippocampal;pheno15,left.paracentral;pheno16,left.pars.opercularis;pheno17,left.pars.orbitalis;pheno18,left.pars.triangularis;pheno19,left.pericalcarine;pheno20,left.postcentral;pheno21,left.posterior.cingulate;pheno22,left.precentral;pheno23,left.precuneus;pheno24,left.rostral.anterior.cingulate;pheno25,left.rostral.middle.frontal;pheno26,left.superior.frontal;pheno27,left.superior.parietal;pheno28,left.superior.temporal;pheno29,left.supramarginal;pheno30,left.transverse.temporal;pheno31,left.insula;pheno32,right.caudal.anterior.cingulate;pheno33,right.caudal.middle.frontal;pheno34,right.cuneus;pheno35,right.entorhinal;pheno36,right.fusiform;pheno37,right.inferior.parietal;pheno38,right.inferior.temporal;pheno39,right.isthmus.cingulate;pheno40,right.lateral.occipital;pheno41,right.lateral.orbitofrontal;pheno42,right.lingual;pheno43,right.medial.orbitofrontal;pheno44,right.middle.temporal;pheno45,right.parahippocampal;pheno46,right.paracentral;pheno47,right.pars.opercularis;pheno48,right.pars.orbitalis;pheno49,right.pars.triangularis;pheno50,right.pericalcarine;pheno51,right.postcentral;pheno52,right.posterior.cingulate;pheno53,right.precentral;pheno54,right.precuneus;pheno55,right.rostral.anterior.cingulate;pheno56,right.rostral.middle.frontal;pheno57,right.superior.frontal;pheno58,right.superior.parietal;pheno59,right.superior.temporal;pheno60,right.supramarginal;pheno61,right.transverse.temporal;pheno62,right.insula;pheno63,cerebellar.vermal.lobules.I.V;pheno64,cerebellar.vermal.lobules.VI.VII;pheno65,cerebellar.vermal.lobules.VIII.X;pheno66,left.basal.forebrain;pheno67,right.basal.forebrain;pheno68,Brain.stem;pheno69,CSF;pheno70,X3rd.ventricle;pheno71,X4th.ventricle;pheno72,optic.chiasm;pheno73,left.lateral.ventricle;pheno74,left.inferior.lateral.ventricle;pheno75,left.cerebellum.exterior;pheno76,left.cerebellum.white.matter;pheno77,left.thalamus.proper;pheno78,left.caudate;pheno79,left.putamen;pheno80,left.pallidum;pheno81,left.hippocampus;pheno82,left.amygdala;pheno83,left.accumbens.area;pheno84,left.ventral.DC;pheno85,left.vessel;pheno86,right.lateral.ventricle;pheno87,right.inferior.lateral.ventricle;pheno88,right.cerebellum.exterior;pheno89,right.cerebellum.white.matter;pheno90,right.thalamus.proper;pheno91,right.caudate;pheno92,right.putamen;pheno93,right.pallidum;pheno94,right.hippocampus;pheno95,right.amygdala;pheno96,right.accumbens.area;pheno97,right.ventral.DC;pheno98,right.vessel;pheno99,gray.matter;pheno100,white.matter;pheno101,total.brain.volume;

### Fig. S9. The effects of PGS for psychiatric, behavioral and socioeconomic traits on child behavioral problems at age three years.

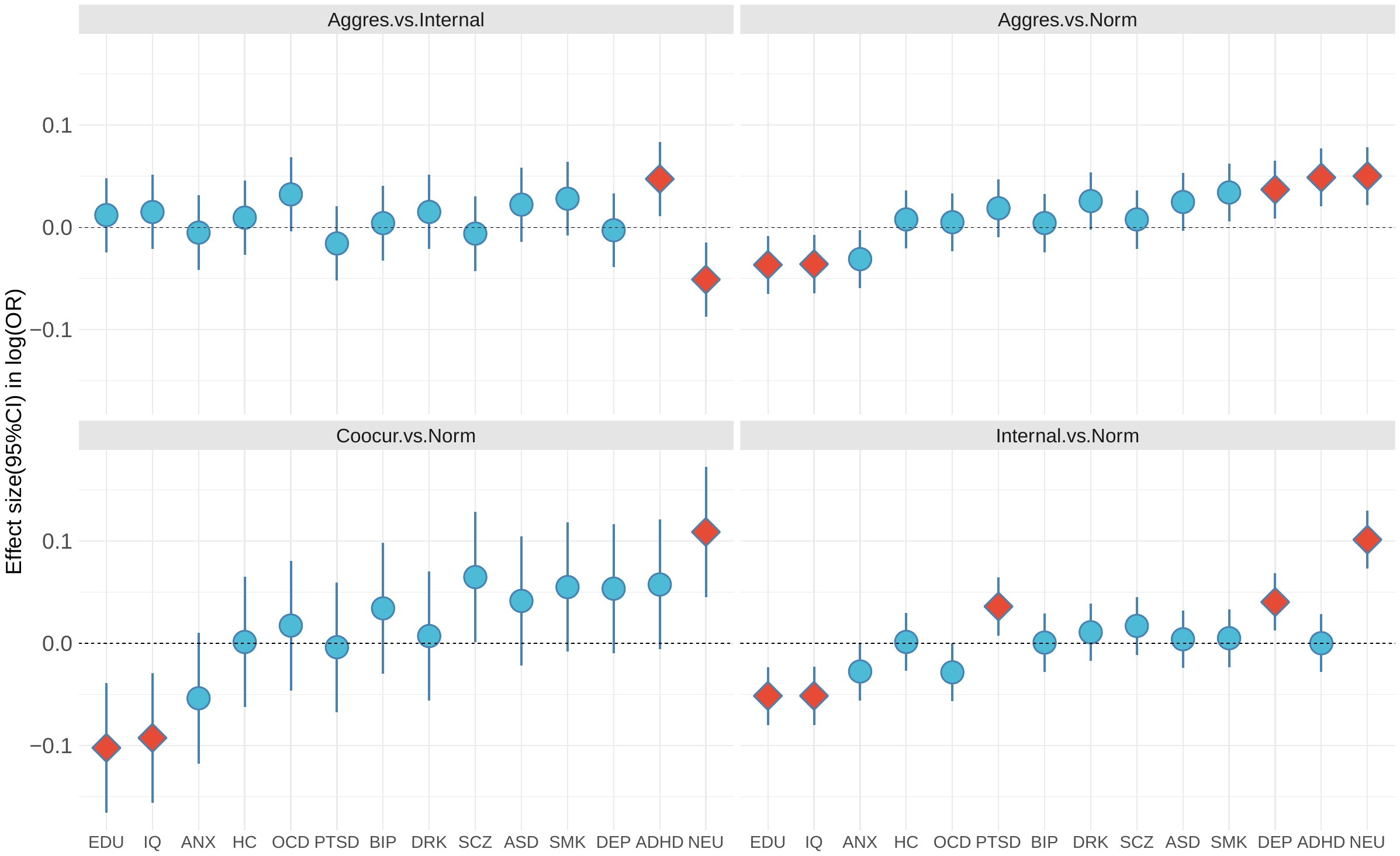

Schizophrenia (SCZ), bipolar disorder (BIP), depression (DEP) , autism spectral disorder (ASD), attention deficit /hyperactivity disorder (ADHD), post-traumatic stress disorder (PTSD), general anxiety (ANX), drinks per week (DRK), smoke per day (SMK), general intelligence (IQ), education attainments (EDU), neuroticism (NEU), Obsessive Compulsive disorder (OCD), and hair color (HC). Statistically significant effects (FDR<0.05) are shown by red diamond; 95% confidence interval are also shown.

### Fig. S10. The effects of PGS for brain structural variation (ROI) on child behavioral problems at age three years.

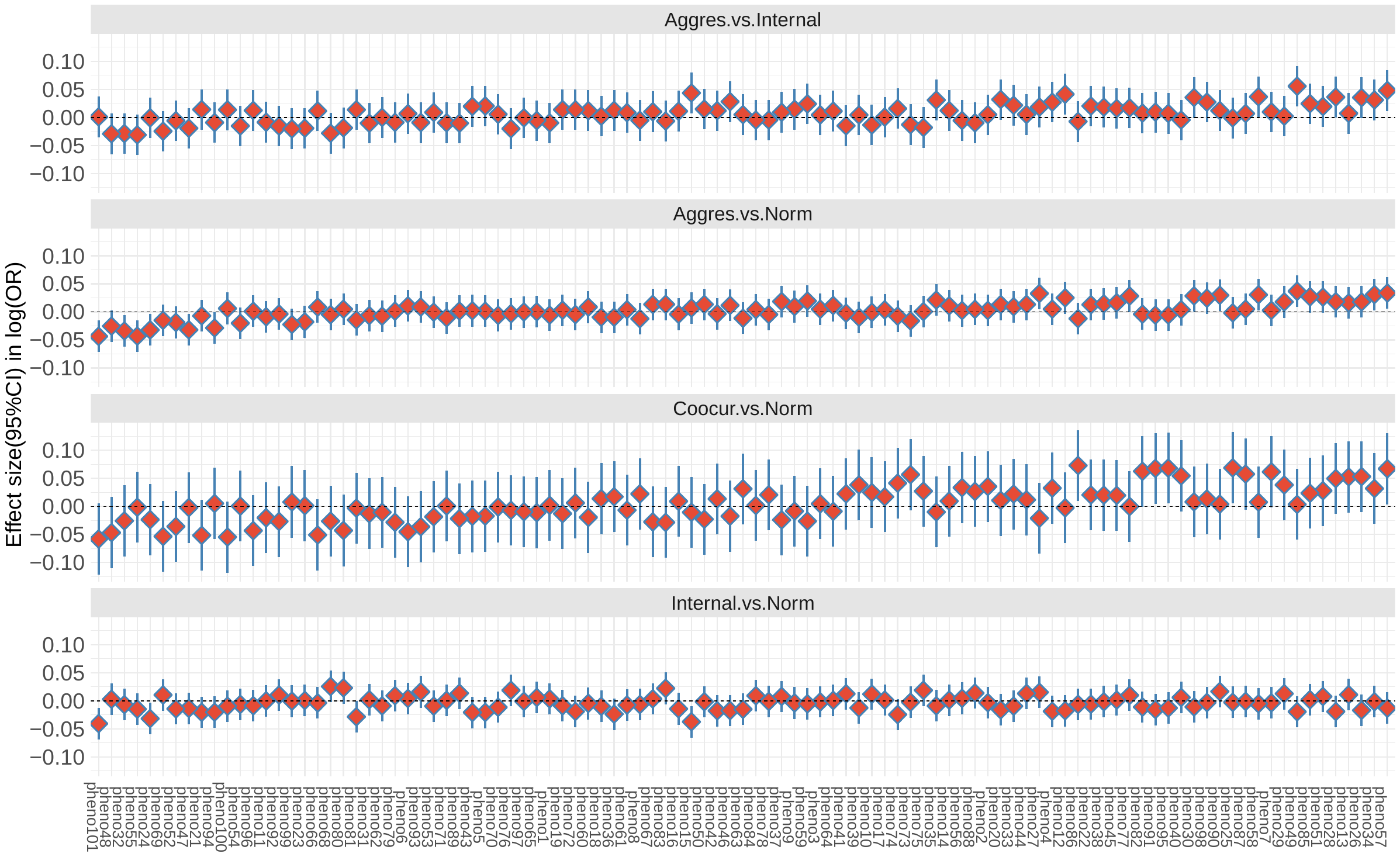

No Statistically significant effects (FDR<0.05) were found; 95% confidence interval are also shown. pheno1,left.caudal.anterior.cingulate;pheno2,left.caudal.middle.frontal;pheno3,left.cuneus;pheno4,left.entorhinal;pheno5,left.fusiform;pheno6,left.inferior.parietal;pheno7,left.inferior.temporal;pheno8,left.isthmus.cingulate;pheno9,left.lateral.occipital;pheno10,left.lateral.orbitofrontal;pheno11,left.lingual;pheno12,left.medial.orbitofrontal;pheno13,left.middle.temporal;pheno14,left.parahippocampal;pheno15,left.paracentral;pheno16,left.pars.opercularis;pheno17,left.pars.orbitalis;pheno18,left.pars.triangularis;pheno19,left.pericalcarine;pheno20,left.postcentral;pheno21,left.posterior.cingulate;pheno22,left.precentral;pheno23,left.precuneus;pheno24,left.rostral.anterior.cingulate;pheno25,left.rostral.middle.frontal;pheno26,left.superior.frontal;pheno27,left.superior.parietal;pheno28,left.superior.temporal;pheno29,left.supramarginal;pheno30,left.transverse.temporal;pheno31,left.insula;pheno32,right.caudal.anterior.cingulate;pheno33,right.caudal.middle.frontal;pheno34,right.cuneus;pheno35,right.entorhinal;pheno36,right.fusiform;pheno37,right.inferior.parietal;pheno38,right.inferior.temporal;pheno39,right.isthmus.cingulate;pheno40,right.lateral.occipital;pheno41,right.lateral.orbitofrontal;pheno42,right.lingual;pheno43,right.medial.orbitofrontal;pheno44,right.middle.temporal;pheno45,right.parahippocampal;pheno46,right.paracentral;pheno47,right.pars.opercularis;pheno48,right.pars.orbitalis;pheno49,right.pars.triangularis;pheno50,right.pericalcarine;pheno51,right.postcentral;pheno52,right.posterior.cingulate;pheno53,right.precentral;pheno54,right.precuneus;pheno55,right.rostral.anterior.cingulate;pheno56,right.rostral.middle.frontal;pheno57,right.superior.frontal;pheno58,right.superior.parietal;pheno59,right.superior.temporal;pheno60,right.supramarginal;pheno61,right.transverse.temporal;pheno62,right.insula;pheno63,cerebellar.vermal.lobules.I.V;pheno64,cerebellar.vermal.lobules.VI.VII;pheno65,cerebellar.vermal.lobules.VIII.X;pheno66,left.basal.forebrain;pheno67,right.basal.forebrain;pheno68,Brain.stem;pheno69,CSF;pheno70,X3rd.ventricle;pheno71,X4th.ventricle;pheno72,optic.chiasm;pheno73,left.lateral.ventricle;pheno74,left.inferior.lateral.ventricle;pheno75,left.cerebellum.exterior;pheno76,left.cerebellum.white.matter;pheno77,left.thalamus.proper;pheno78,left.caudate;pheno79,left.putamen;pheno80,left.pallidum;pheno81,left.hippocampus;pheno82,left.amygdala;pheno83,left.accumbens.area;pheno84,left.ventral.DC;pheno85,left.vessel;pheno86,right.lateral.ventricle;pheno87,right.inferior.lateral.ventricle;pheno88,right.cerebellum.exterior;pheno89,right.cerebellum.white.matter;pheno90,right.thalamus.proper;pheno91,right.caudate;pheno92,right.putamen;pheno93,right.pallidum;pheno94,right.hippocampus;pheno95,right.amygdala;pheno96,right.accumbens.area;pheno97,right.ventral.DC;pheno98,right.vessel;pheno99,gray.matter;pheno100,white.matter;pheno101,total.brain.volume;

### Fig. S11. The effects of PGS for psychiatric, behavioral and socioeconomic traits on child behavioral problems at age five years.

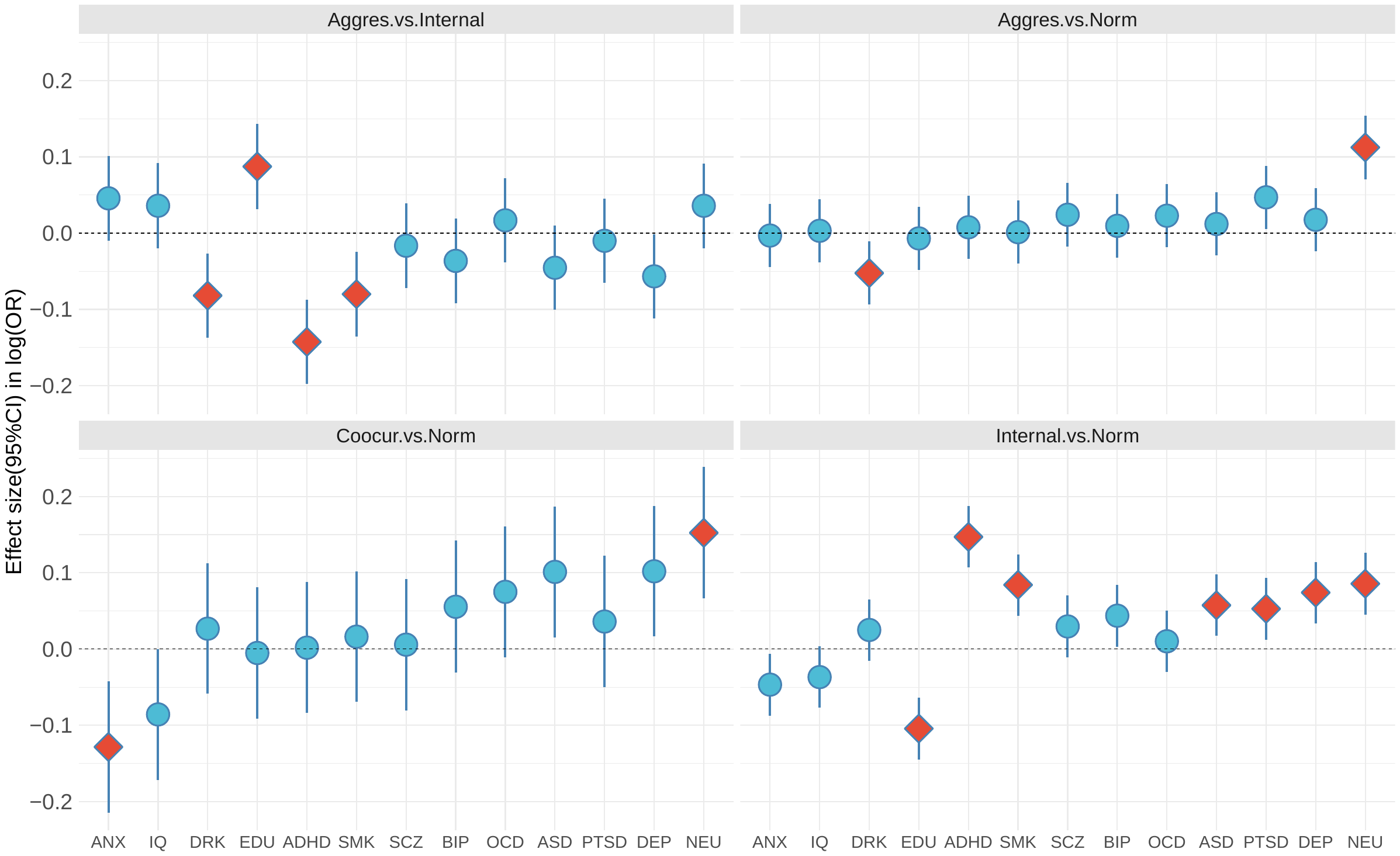

Schizophrenia (SCZ), bipolar disorder (BIP), depression (DEP) , autism spectral disorder (ASD), attention deficit /hyperactivity disorder (ADHD), post-traumatic stress disorder (PTSD), general anxiety (ANX), drinks per week (DRK), smoke per day (SMK), general intelligence (IQ), education attainments (EDU), neuroticism (NEU), Obsessive Compulsive disorder (OCD), and hair color (HC). Statistically significant effects (FDR<0.05) are shown by red diamond; 95% confidence interval are also shown.

### Fig. S12. The effects of PGS for brain structural variation (ROI) on child behavioral problems at age five years

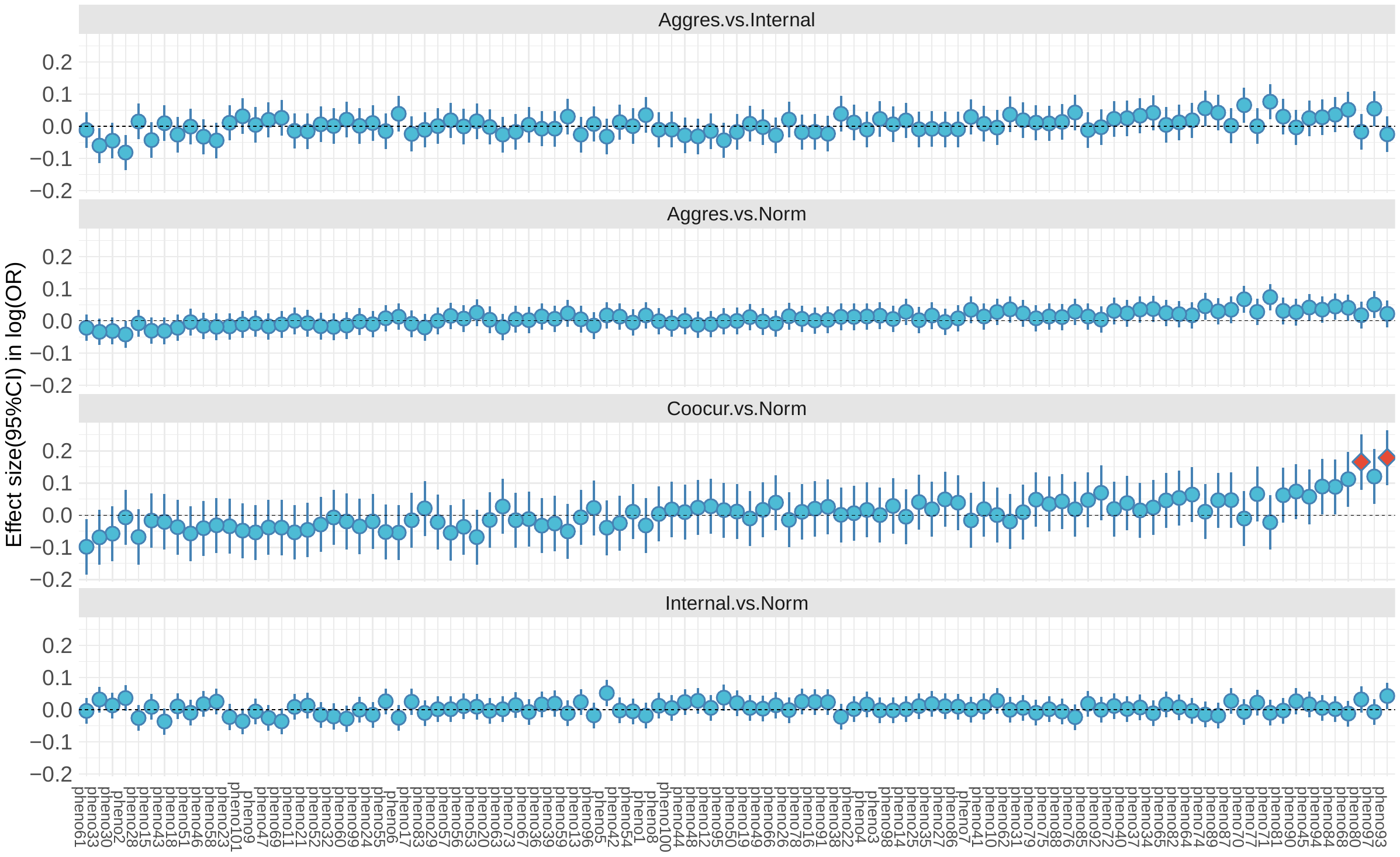

Statistically significant effects (FDR<0.05) are shown by red diamond; the 95% confidence interval are also shown. pheno1,left.caudal.anterior.cingulate;pheno2,left.caudal.middle.frontal;pheno3,left.cuneus;pheno4,left.entorhinal;pheno5,left.fusiform;pheno6,left.inferior.parietal;pheno7,left.inferior.temporal;pheno8,left.isthmus.cingulate;pheno9,left.lateral.occipital;pheno10,left.lateral.orbitofrontal;pheno11,left.lingual;pheno12,left.medial.orbitofrontal;pheno13,left.middle.temporal;pheno14,left.parahippocampal;pheno15,left.paracentral;pheno16,left.pars.opercularis;pheno17,left.pars.orbitalis;pheno18,left.pars.triangularis;pheno19,left.pericalcarine;pheno20,left.postcentral;pheno21,left.posterior.cingulate;pheno22,left.precentral;pheno23,left.precuneus;pheno24,left.rostral.anterior.cingulate;pheno25,left.rostral.middle.frontal;pheno26,left.superior.frontal;pheno27,left.superior.parietal;pheno28,left.superior.temporal;pheno29,left.supramarginal;pheno30,left.transverse.temporal;pheno31,left.insula;pheno32,right.caudal.anterior.cingulate;pheno33,right.caudal.middle.frontal;pheno34,right.cuneus;pheno35,right.entorhinal;pheno36,right.fusiform;pheno37,right.inferior.parietal;pheno38,right.inferior.temporal;pheno39,right.isthmus.cingulate;pheno40,right.lateral.occipital;pheno41,right.lateral.orbitofrontal;pheno42,right.lingual;pheno43,right.medial.orbitofrontal;pheno44,right.middle.temporal;pheno45,right.parahippocampal;pheno46,right.paracentral;pheno47,right.pars.opercularis;pheno48,right.pars.orbitalis;pheno49,right.pars.triangularis;pheno50,right.pericalcarine;pheno51,right.postcentral;pheno52,right.posterior.cingulate;pheno53,right.precentral;pheno54,right.precuneus;pheno55,right.rostral.anterior.cingulate;pheno56,right.rostral.middle.frontal;pheno57,right.superior.frontal;pheno58,right.superior.parietal;pheno59,right.superior.temporal;pheno60,right.supramarginal;pheno61,right.transverse.temporal;pheno62,right.insula;pheno63,cerebellar.vermal.lobules.I.V;pheno64,cerebellar.vermal.lobules.VI.VII;pheno65,cerebellar.vermal.lobules.VIII.X;pheno66,left.basal.forebrain;pheno67,right.basal.forebrain;pheno68,Brain.stem;pheno69,CSF;pheno70,X3rd.ventricle;pheno71,X4th.ventricle;pheno72,optic.chiasm;pheno73,left.lateral.ventricle;pheno74,left.inferior.lateral.ventricle;pheno75,left.cerebellum.exterior;pheno76,left.cerebellum.white.matter;pheno77,left.thalamus.proper;pheno78,left.caudate;pheno79,left.putamen;pheno80,left.pallidum;pheno81,left.hippocampus;pheno82,left.amygdala;pheno83,left.accumbens.area;pheno84,left.ventral.DC;pheno85,left.vessel;pheno86,right.lateral.ventricle;pheno87,right.inferior.lateral.ventricle;pheno88,right.cerebellum.exterior;pheno89,right.cerebellum.white.matter;pheno90,right.thalamus.proper;pheno91,right.caudate;pheno92,right.putamen;pheno93,right.pallidum;pheno94,right.hippocampus;pheno95,right.amygdala;pheno96,right.accumbens.area;pheno97,right.ventral.DC;pheno98,right.vessel;pheno99,gray.matter;pheno100,white.matter;pheno101,total.brain.volume;

### Fig. S13. Distributions of PGS for traits shown significant effects on child behavioral problems.

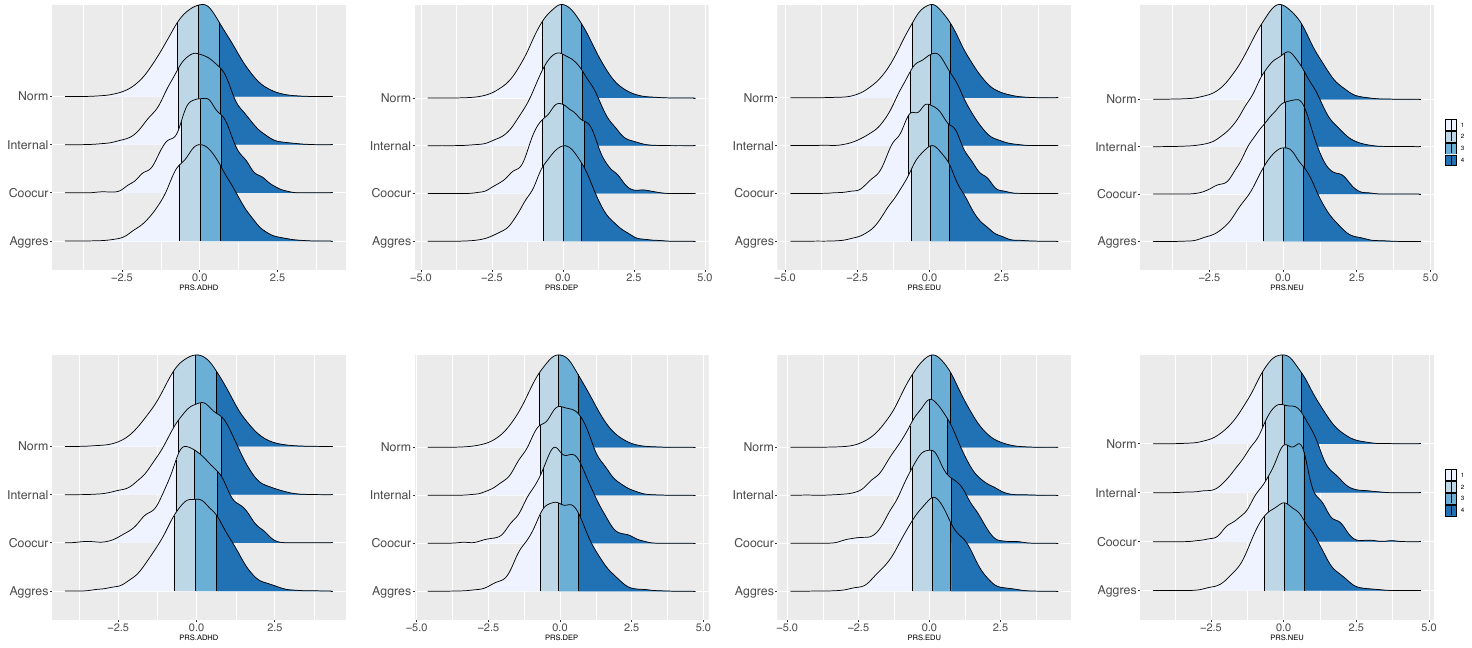

First row: age at three years; second row: age at five years; Colors indicate the four quantiles.

### Fig. S14. The Manhattan plots for EWAS of child behavioral problems measure at age three years.

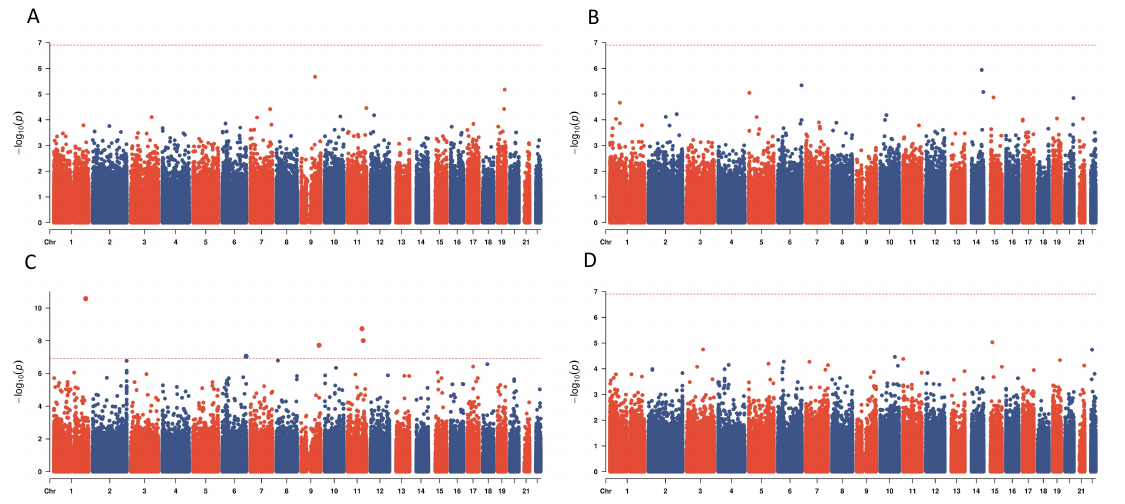

A. Aggressive vs. internalizing behavior; B. aggressive vs. normal behavior; C. co-occurring vs. normal behavior; D. internalizing vs. normal behavior.

### Fig. S15. The QQ plots for EWAS of child behavioral problems measure at age three years.

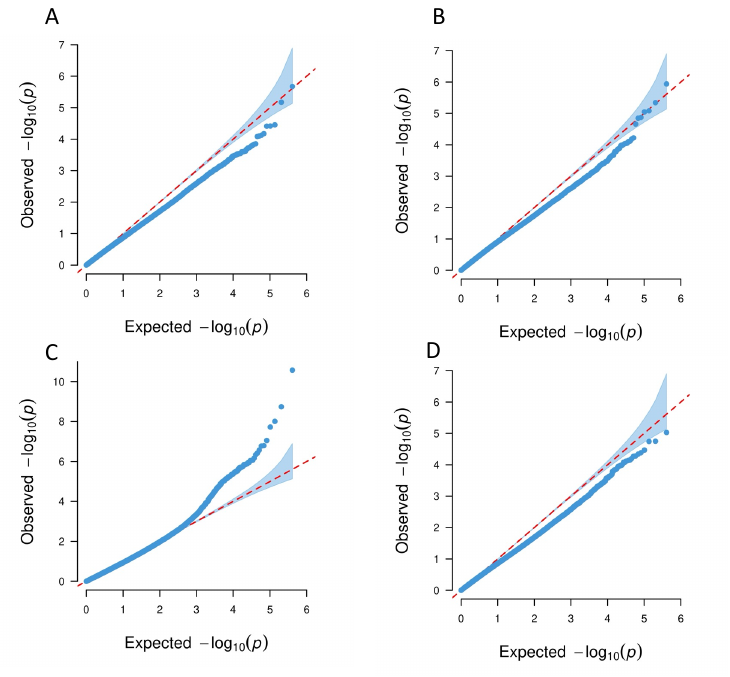

A. Aggressive vs. internalizing behavior; B. aggressive vs. normal behavior; C. co-occurring vs. normal behavior; D. internalizing vs. normal behavior.

### Fig. S16. The volcano plots for EWAS of child behavioral problems measure at age three years.

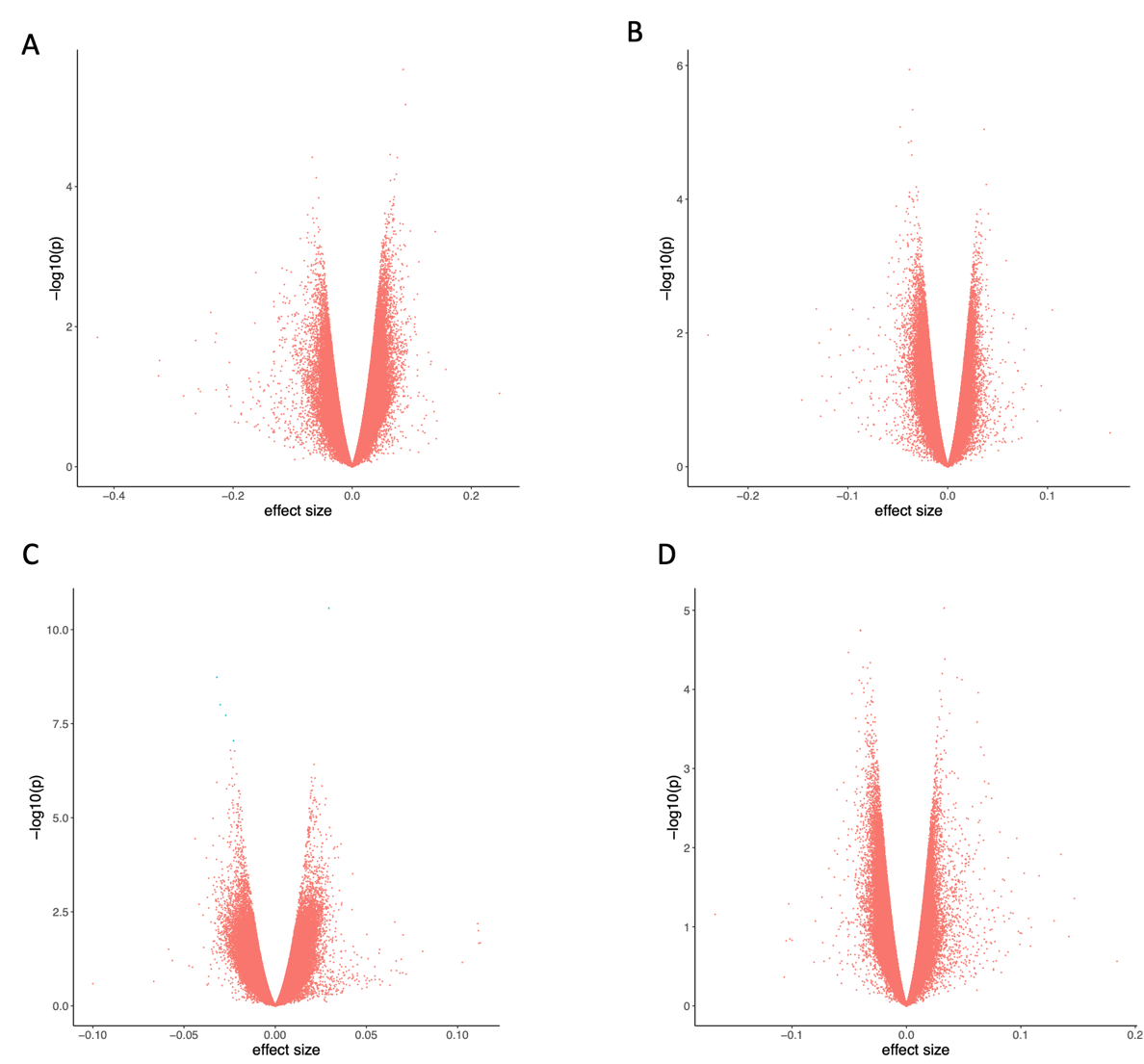

A. Aggressive vs. internalizing behavior; B. aggressive vs. normal behavior; C. co-occurring vs. normal behavior; D. internalizing vs. normal behavior.

### Fig. S17. The Manhattan plots for EWAS of child behavioral problems measure at age five years.

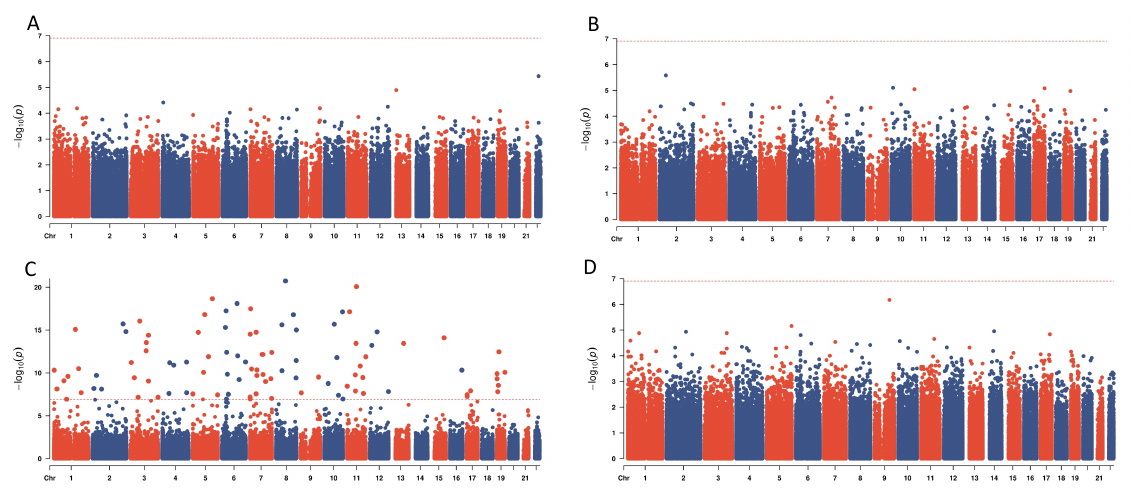

A. Aggressive vs. internalizing behavior; B. aggressive vs. normal behavior; C. co-occurring vs. normal behavior; D. internalizing vs. normal behavior.

### Fig. S18. The QQ plots for EWAS of child behavioral problems measure at age five years.

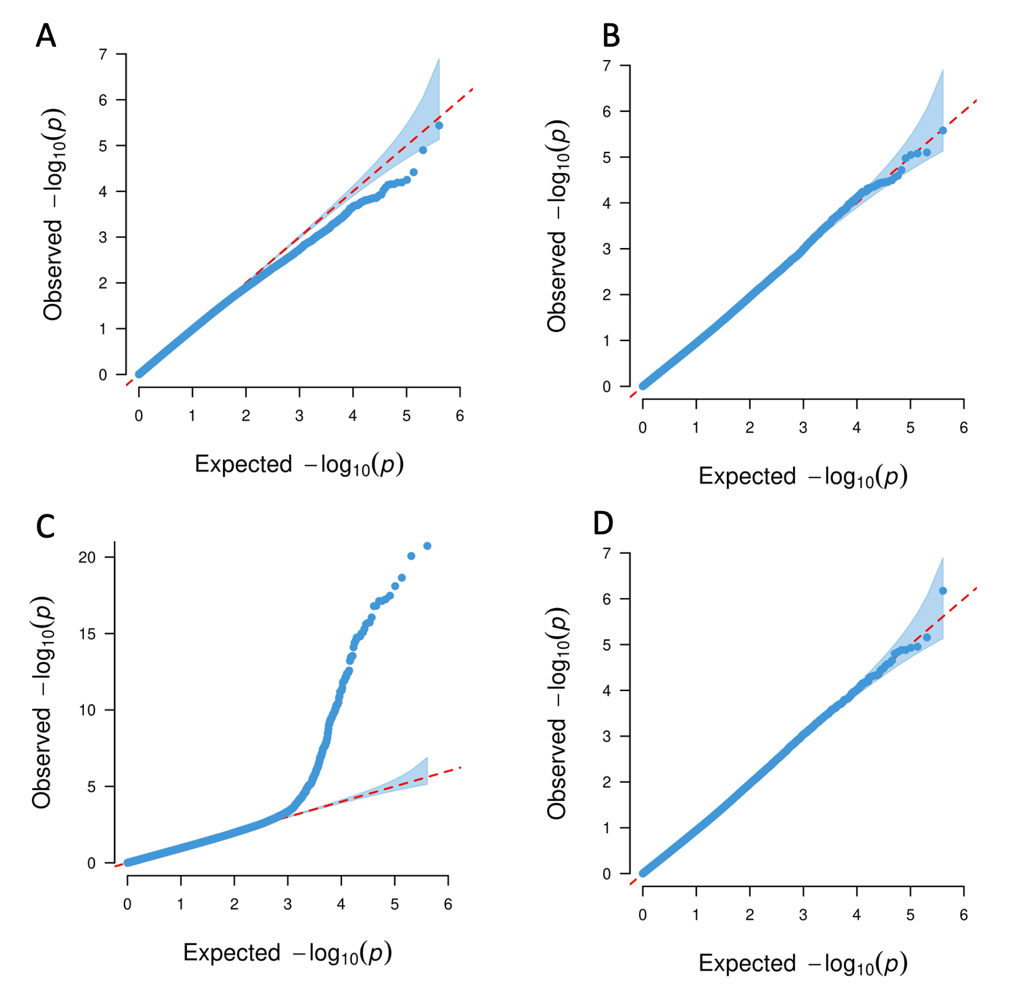

A. Aggressive vs. internalizing behavior; B. aggressive vs. normal behavior; C. co-occurring vs. normal behavior; D. internalizing vs. normal behavior.

### Fig. S19. The volcano plots for EWAS of child behavioral problems measure at age five years.

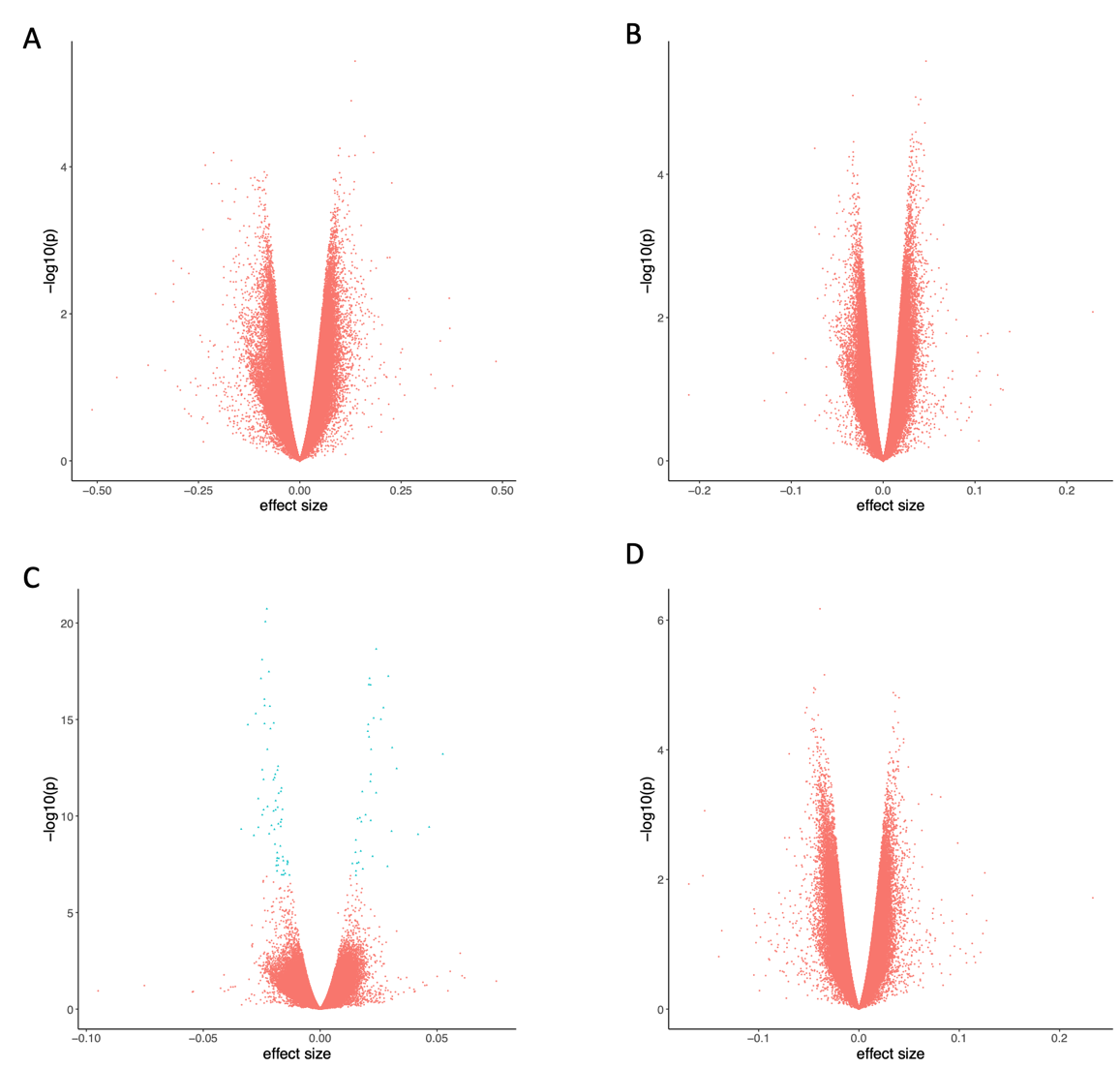

A. Aggressive vs. internalizing behavior; B. aggressive vs. normal behavior; C. co-occurring vs. normal behavior; D. internalizing vs. normal behavior.

### Fig. S20. Gene clusters identified by GeneNetwork for the genes from EWAS of Co-occuring vs normal behavior.

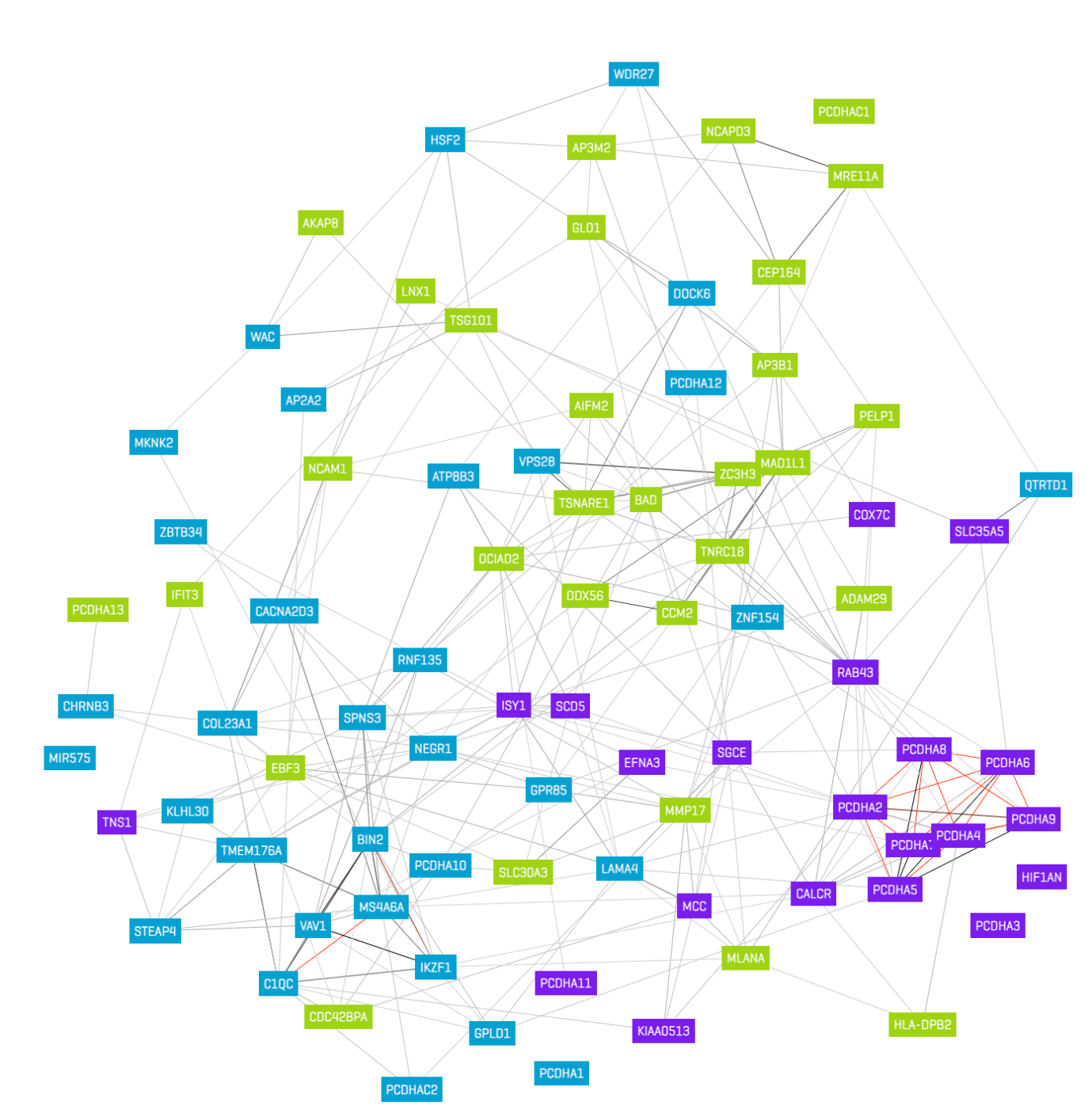

### Fig. S21. Gene set enrichment for the Reactome database for the gene clusters from Fig. S20.

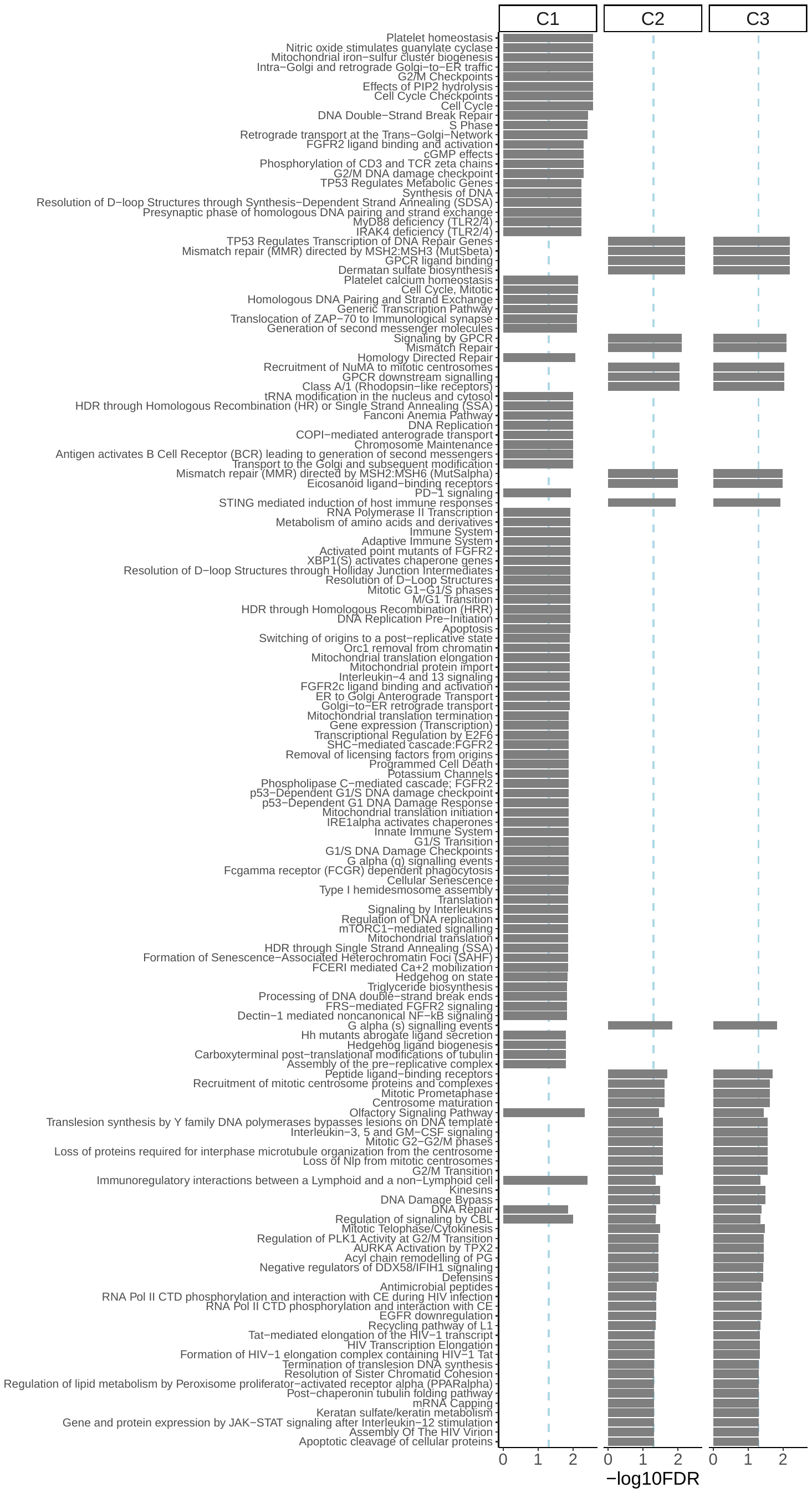

### Fig. S22 Gene set enrichment for the Reactome database for the genes in the black module.

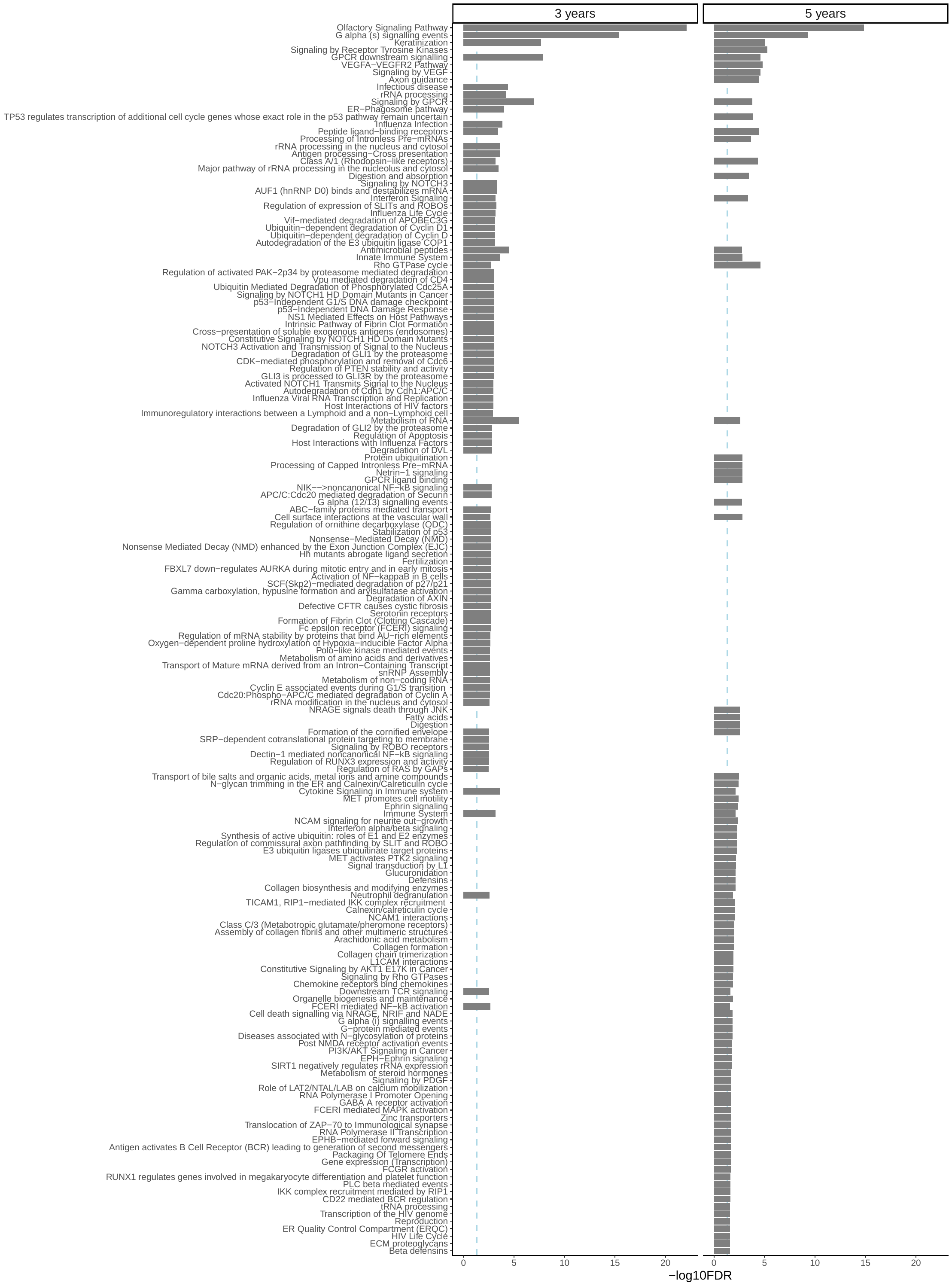

### Fig. S23. Levels of internalizing problems and aggression per profile at age 3 years.

Age: 3 years

Z-scores

|  | Internalizing problems Z-score | Aggession Z-score |
| --- | --- | --- |
| Low level profile | -0.49 | -0.42 |
| Primarily Aggressive profile | 0.25 | 1.52 |
| Primarily Internalizing profile | 1.38 | 0.07 |
| Co-occurrent profile | 2.99 | 1.56 |

### Fig. S24. Levels of internalizing problems and aggression per profile at age 5 years.

Age: 5 years

Z-scores

|  | Internalizing problems Z-score | Aggession Z-score |
| --- | --- | --- |
| Low level profile | -0.34 | -0.31 |
| Primarily Aggressive profile | 0.2 | 2 |
| Primarily Internalizing profile | 1.73 | 0.06 |
| Co-occurrent profile | 3.66 | 1.62 |

### Fig. S25. Power plot for the WGNCA analysis for DNAm at age 3 years..

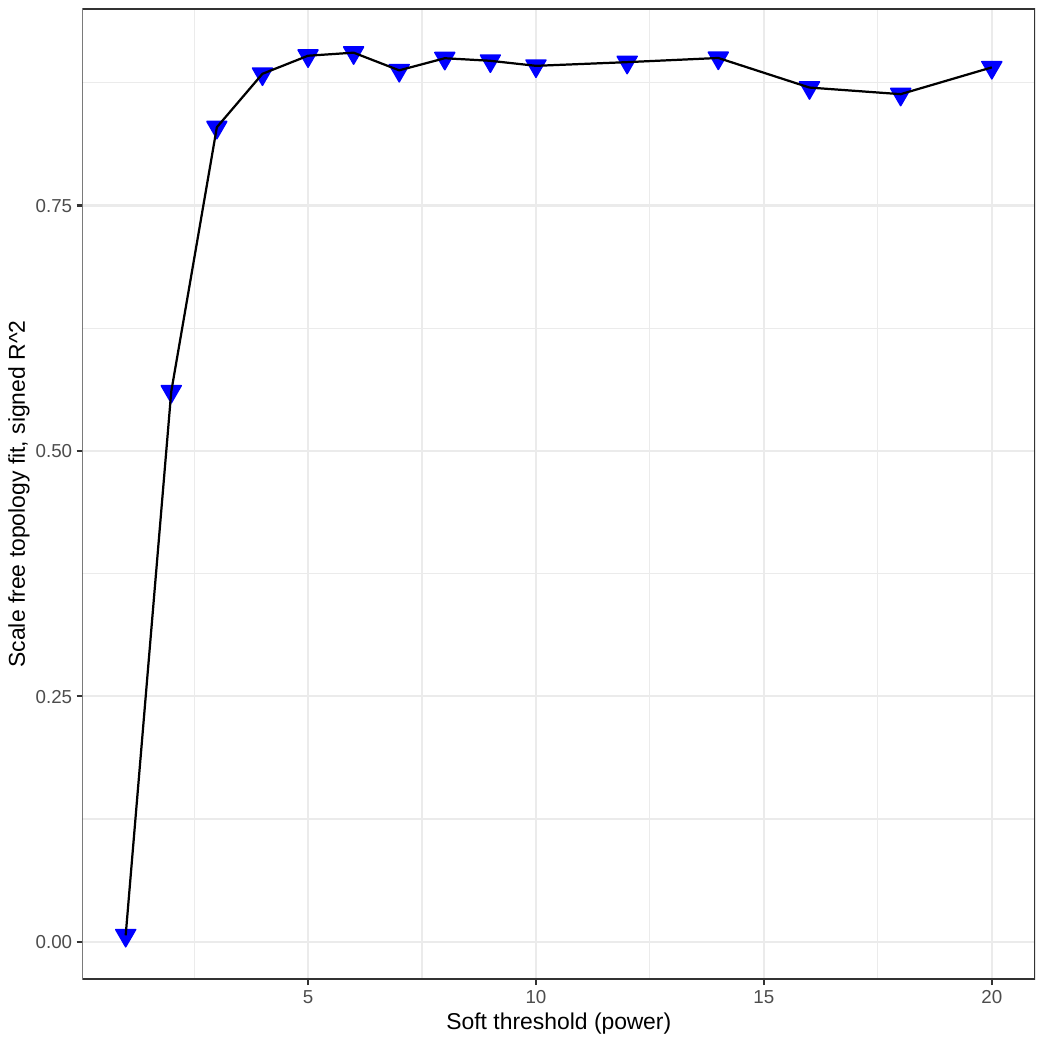

### Fig. S26. Power plot for the WGNCA analysis for DNAm at age 5 years.

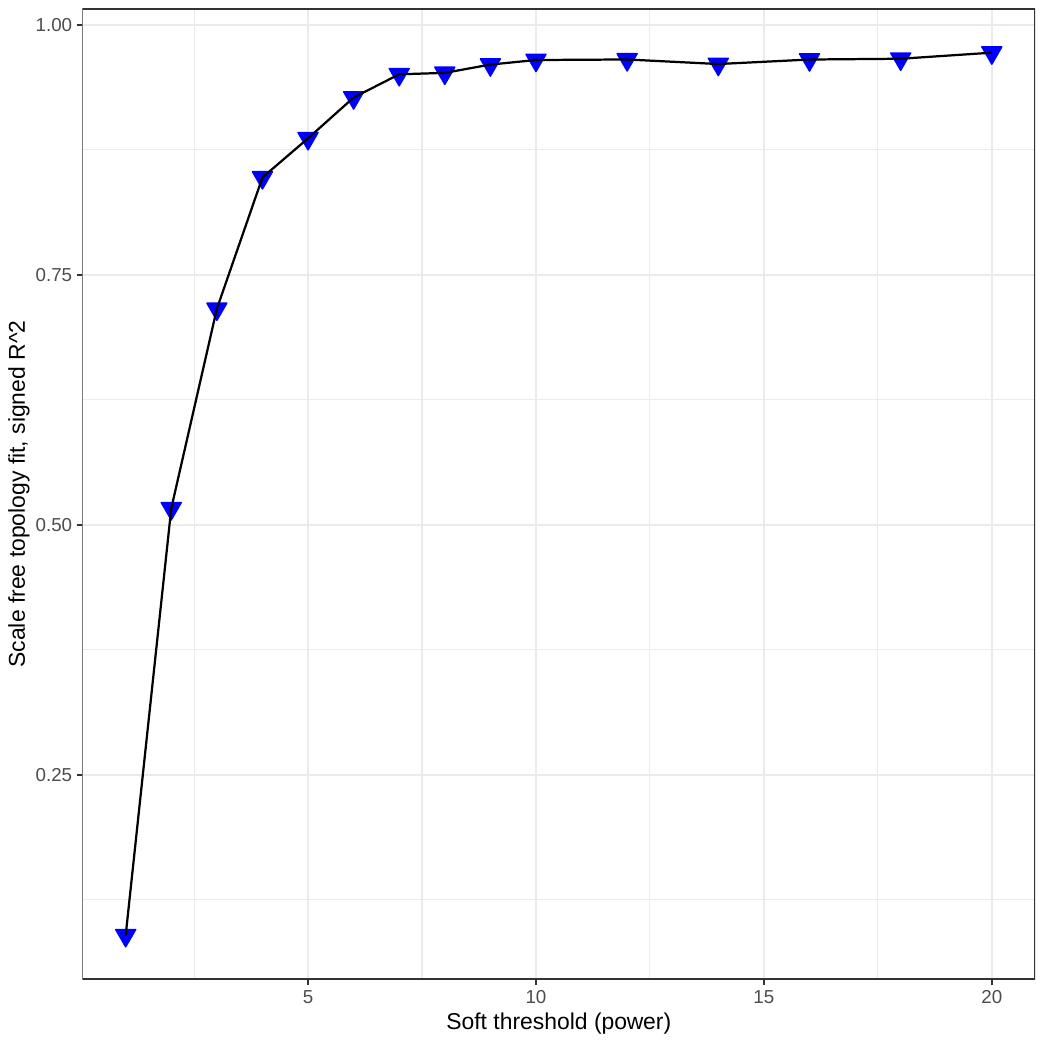
